# Pro-inflammatory innate-like T cells are expanded in the blood and inflamed intestine in Crohn’s Disease

**DOI:** 10.1101/2022.05.12.22274864

**Authors:** Cristina M. Chiarolla, Axel R. Schulz, Michael Meir, Simone Reu-Hofer, Addi J. Romero-Olmedo, Michael Lohoff, Andreas Rosenwald, Hyun-Dong Chang, Nicolas Schlegel, Henrik E. Mei, Friederike Berberich-Siebelt

**Author notes:** Address correspondence to: Dr. Friederike Berberich-Siebelt, Institute of Pathology, University of Würzburg, Josef-Schneider-Str. 2, 97080 Wuerzburg, ORCID profile: 0000-0003-1673-4175.

## Abstract

A complex and tissue-specific network of cells including T lymphocytes maintains intestinal homeostasis. To address disease and tissue-specific alterations, we performed a T cell-centric mass cytometry analysis of peripheral and intestinal lymphocytes from patients with Crohn’s disease (CD) and healthy donor PBMCs. We compared inflamed and not inflamed tissue areas of bowel resections. Chronic inflammation enforced activation, exhaustion and terminal differentiation of CD4^+^ and CD8^+^ T cells and an enrichment of CD4^+^Foxp3^+^ cells (Tregs) in inflamed intestine. However, tissue-repairing Tregs decreased, while enigmatic rare Foxp3^+^ T-cell subsets appeared upon inflammation. In vitro assays revealed that those subsets, e.g. CD4^+^Foxp3^+^HLA-DR^+^TIGIT^−^ and CD4^+^Foxp3^+^CD56^+^, express pro-inflammatory IFN-γ. Some T-conventional (Tcon) cells tended towards innateness. In blood of CD patients, not well studied CD4^+^ and CD8^+^ subsets of CD16^+^CCR6^+^CD127^+^ T cells appeared anew, a phenotype reproducible by incubation of healthy blood T cells with patient blood plasma. Together, these findings suggest a bias towards innate-like pro-inflammatory Tregs and innate-like Tcon, which act with less specific cytotoxicity. Most likely, this is both cause and consequence of intestinal inflammation during CD.

**Graphical Abstract:** 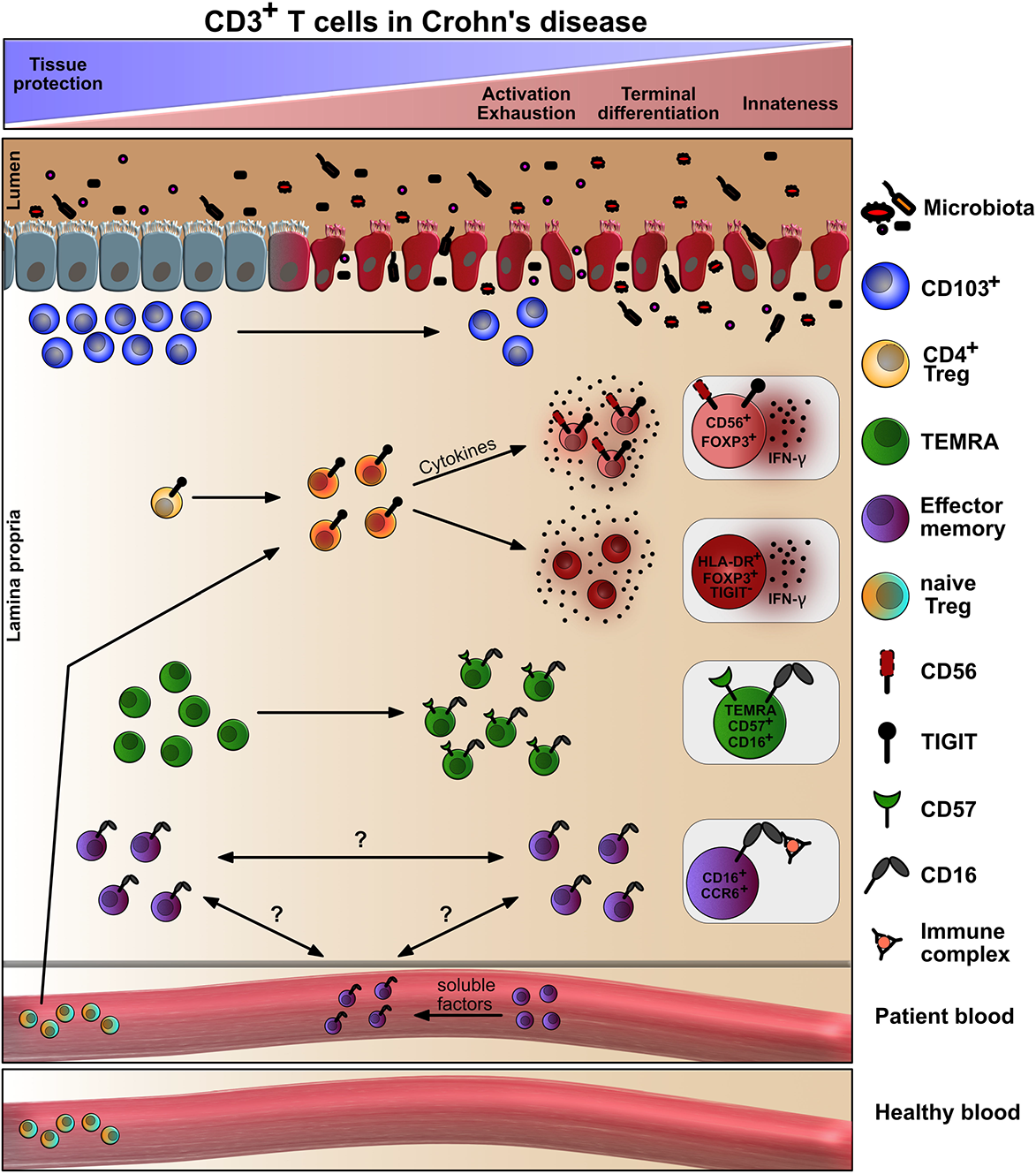

## Introduction

Maintenance of intestinal homeostasis is dependent upon the immune system’s ability to remain tolerant to environmental antigens and commensals while mounting appropriate immune responses to pathogens that utilize the gut as a primary site of entry. A breakdown of tolerance contributes to inflammatory bowel disease (IBD) comprising two major types that are known as Crohn’s disease (CD) and ulcerative colitis (UC). Despite extensive research, the pathophysiology underlying IBD remains unclear. Current concepts suggest that IBD is caused by multiple factors including genetic predisposition, aberrant immune reactions, altered microbiota, environmental factors and loss of intestinal epithelial barrier function (1, 2).

Patients with IBD suffer from abdominal cramping, diarrhea, rectal bleeding, and weight loss due to local intestinal dysfunction. In addition, these patients are threatened by long-term complications including increased susceptibility to colorectal cancer and extra-intestinal manifestations such as arthritis. A significant number of CD patients – characterized by transmural granulomatous reactions – require surgery because of refractory e.g. fistulizing or stenotic disease (3).

Since the aberrant immune response in IBD is idiopathic, corticosteroids and broad immunosuppressants are widely used drugs (2). For longterm, pro-inflammatory cytokines – mostly secreted by pro-inflammatory T cells – are targeted predominantly by anti-TNF biologicals or JAK inhibitors. However, to elicit changes in lymphocyte subsets of patients with inflammatory diseases, often blood samples are analyzed in comparison to healthy controls. This can be very informative and describe cell markers or subpopulations which are indicative for disease. However, how well tissue-based immunopathology is reflected by blood analyses is continuously discussed (4). Direct analysis of cells isolated from the inflamed tissues promises more immediate information, for example whether an underrepresented subpopulation among PBMCs migrates to the tissue to be enriched there or whether this subpopulation is overall lessened upon disease onset. One prominent population often evaluated in peripheral blood are T-regulatory cells (Tregs) under the claim that they are diminished in inflammatory diseases. Indeed, it has been observed early on that CD4^+^CD25^hi^ Tregs are less among PBMCs, although the inflamed mucosa of IBD patients then proved to be enriched in Tregs (5).

To assess frequencies and phenotypes of Tregs is crucial, because they elicit self-tolerance and long-term immune homeostasis, protect from tissue inflammation and exert tissue repair by a variety of mechanisms (6). Tregs – CD4^+^, but also CD8^+^ T cells – express the high-affinity IL-2 receptor α-chain (CD25) and the transcription factor Forkhead box P-3 (FOXP3) which is essential for their development, stability and suppressive function (7–9). Upon T-cell receptor (TCR) engagement, CD25 and, at least in humans, FOXP3 are also upregulated by T-conventional (Tcon) cells. In human Tcon, FOXP3 expression is low and transient. Since pronounced IL-7Rα/CD127 is exclusively found on Tcon, the CD4^+^CD25^hi^CD127^lo/–^FOXP3^hi^ phenotype defines the majority of Tregs (10, 11).

Beyond naturally occurring thymus-derived tTregs, FOXP3^hi^ Tregs can stem from naive Tcon in the periphery (pTregs), where their induction depends on TCR engagement in conjunction with cytokine-mediated signals (12, 13). This reflects the micromilieu-dependent lineage commitment – and a certain degree of plasticity – of any peripheral CD4^+^ or CD8^+^ Tcon (14, 15).

Besides their potential to differentiate into specific cytokine-producing T-helper and also T-cytotoxic subtypes, T cells are acknowledged as a heterogeneous pool of effector and memory cells (16, 17). Here, T cells are recognized as naïve (CD45RA^+^ CD27^+^ CCR7^+^), short lived effector (CD127^−^ KLRG1^+^ SLEC), memory precursor effector (CD127^+^KLRG1^−^MPEC), central (CD45RA^−^ CD127^+^CD27^+^CCR7^+^ T_CM_), effector (CD45RA^−^CD127^+^CD27^−^CCR7^−^ T_EM_) and peripheral (CD45RA^−^CD127^+^CD27^+^CCR7^−^ T_PM_) memory cells along with a terminally differentiated subset that re-expresses CD45RA (CD45RA^+^CD127^−^CD27^−^CCR7^−^ T_EMRA_) and accumulates upon chronic infections. Data obtained from organ donors revealed large populations of tissue-resident T_RM_ cells and tissue-specific frequencies of T_CM_, T_EM_ and T_EMRA_, which were – again – not reflective of subset representation within the circulation (18).

Broad immune cell phenotyping of PBMCs vs. tissues of IBD patients has been performed (19–21). Since CD3^+^ T cells, both CD4^+^ and CD8^+^, are thought to be key players in CD (22, 23), we here aimed to perform a deep immune characterization using mass cytometry (Cytometry by Time-of-Flight, CyTOF) together with manual and computation data analysis, and offer a detailed description of CD3^+^ T-cell heterogeneity in a CD context. Moreover, to relate subtypes stringently to areas of inflammation, we further discriminated between inflamed and not inflamed lamina propria (LP) as well as epithelium of the same CD patient, additionally analyzing their blood in comparison to matched healthy donors (HD) (Figure 1A). We found loss of tissue-protective T cells and observed a tendency towards activation, exhaustion and terminal differentiation for both CD4^+^ and CD8^+^ T cells. Intriguingly, they acquired innate-like effector phenotypes. The upregulation of proteins known from innate immune cells was paralleled by a partly pro-inflammatory phenotype of Tregs accumulating in inflamed areas. Finally, we noticed the emergence of circulating innate-like CD16^+^CCR6^+^ T cells, in vitro inducible by CD patients’ plasma and prone to serve as an easily accessible disease marker in future.

**Figure 1.**
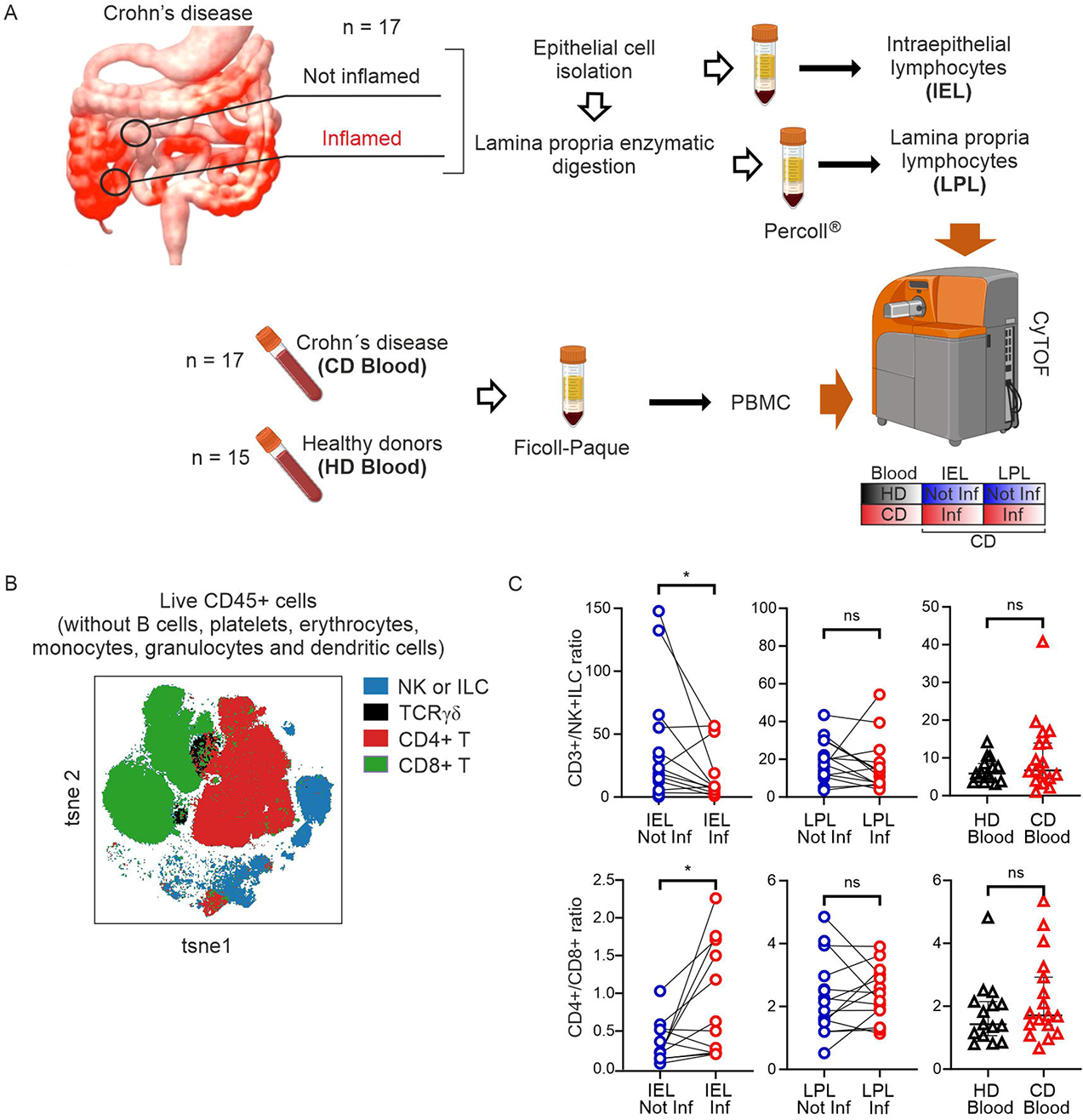
Global mass cytometric characterization of CD45^+^ cells isolated from CD patients’ intestinal and blood samples in comparison with HDs’ blood. (**A**) (Created with BioRender.com.) Experimental setup for mass cytometric analyses of lymphocytes from inflamed and not inflamed intestinal tissue of CD patients alongside with matched PBMC and HD PBMC (CD, n=17; HD, n=15). (**B**) tsne plot depicting live CD45^+^ cells from all samples after removal of unwanted cells as shown in Figure S1A. Colors indicate 5 major CD45^+^ cell populations identified by manual gating in Figure S1A. Samples with rather few cell events were excluded from the data analysis (final analysis was carried out on n=16 IEL Not Inf; n=12 IEL Inf; n=16 LPL Not Inf; n=14 LPL Inf, n=16 CD blood, n=15 HD blood). (**C**) Comparison of CD3^+^ T cell / NK+ILC cell ratio (top) and CD4^+^/CD8^+^ T-cell ratio (bottom) in inflamed vs. not inflamed epithelium and LP and CD vs. HD blood. Each data point represents one patient. IEL, n=12; LPL, n=14; CD blood, n=17; HD blood, n=15. Statistical significance was calculated using two-tailed Wilcoxon test for paired intestinal samples and Mann-Whitney test for the blood samples; *, p<0.05.

## Results

### The frequency of CD8^+^ T cells is diminished in the inflamed epithelium of CD patients

To determine the phenotype of CD3^+^ T-cell subsets within the intestinal tissues of CD patients (Supplemental Table 1 and 2) in comparison to their blood, we chose *Cytometry by time of flight* (CyTOF). By the use of heavy metal isotope-conjugated monoclonal antibodies, we simultaneously quantified 42 different intracellular and surface markers on individual cells (Supplemental Table 3)(24). We manually gated CD4^+^, CD8^+^, CD4^−^CD8^−^ (DN), CD4^+^CD8^+^ (DP), and TCRγδ^+^ T cells as well as MAIT and CD3^−^ NK cells together with innate lymphoid cells (ILC) from live CD45^+^ cells (Supplemental Figure 1A). When CD45^+^ cells from all the samples – PBMCs, IELs and LPLs from inflamed (Inf) and not inflamed (Not Inf) tissues of 17 CD patients as well as PBMCs from 15 volunteers – were visualized by a bidimensional tSNE plot, the manually pre-gated populations appeared clearly separated (Figure 1B).

Next, we determined if there were differences in frequencies of those pre-gated major cell populations between inflamed vs. not inflamed tissues or between CD patients’ and HD blood. Interestingly, although DN CD3^+^ T cells were less abundant among PBMCs of CD patients in comparison to HD, they were enriched in the LP upon inflammation (Supplemental Figure 1B), consistent with the recruitment or retention of otherwise circulating cells to inflamed tissue. Other populations, i.e. DP and TCRγδ^+^ T cells as well as MAIT cells were unchanged in occurrence upon inflammation within the CD3^+^ lymphocytes at all sites tested (Supplemental Figure1, C-E). However, in inflamed epithelia, although not in LP or blood, the ratio of CD3^+^ T cells to CD3^−^ innate lymphocytes decreased, whereas the ratio of CD4^+^ to CD8^+^ T cells increased significantly (Figure 1C). Overall, patients behaved similarly with respect to subset distribution allowing the correlation of heightened or diminished subset appearance with each other (Supplemental Figure 2). For example, among IELs and LPLs, innate-like NK/ILC, TCRγδ^+^ and MAIT cells positively correlated with CD4^+^ Tregs upon inflammation. In inflamed epithelia this was extendable to DP CD3^+^ T cells, whose abundance in inflamed LP and blood further linked well with the CD8^+^ Treg subset. Together, this indicated a robust relative loss of CD8^+^ Tcon compared to CD4^+^ T cells and especially CD4^+^ Tregs as well as innate-like T cells upon inflammation.

### Protective CD103^+^ *γδ T_RM_ cells* are lost in the inflamed LP of CD patients

To analyze distinct major subsets of T cells in greater depth, we chose a nested approach. After identifying major subsets as shown in Figure 1/S1/S2, we performed a second set of FlowSOM clustering for CD4^+^, CD8^+^, TCRγδ^+^ and MAIT cells separately on the different pre-gated CD3^+^ T-cell populations. CD3^+^ MAIT cells, gated as TCRα7.2^+^ and CD161^+^ (KLRB1), as well as TCRγδ^+^ T cells were each divided in 10 clusters (Supplemental Figure 3A, Figure 2A). Heat maps demonstrate the relative average expression of every protein measured in a given cluster (Supplemental Figure 3B, Figure 2B). To assess possible changes between inflamed and not inflamed tissues, the percentage within the total CD3^+^ population was calculated for each cluster. Quantitative changes are indicated. CD4^+^CD28^+^CD127^+^CXCR3^+^CD103^−^ MAIT_4 cells, which encompassed the largest sub-cluster in LP, were significantly less noticeable in CD patients’ blood than in HD (Supplemental Figure 3B and C), whereas the CD103^+^ TCRγδ_03 was considerably less prominent in inflamed LP. This is the clearly distinct TCR γδ^+^ subtype expressing not only CD103, but also CD8, NKp46 and TIGIT (Figure 2C), described to have anti-inflammatory/immune-regulatory properties (25).

**Figure 2.**
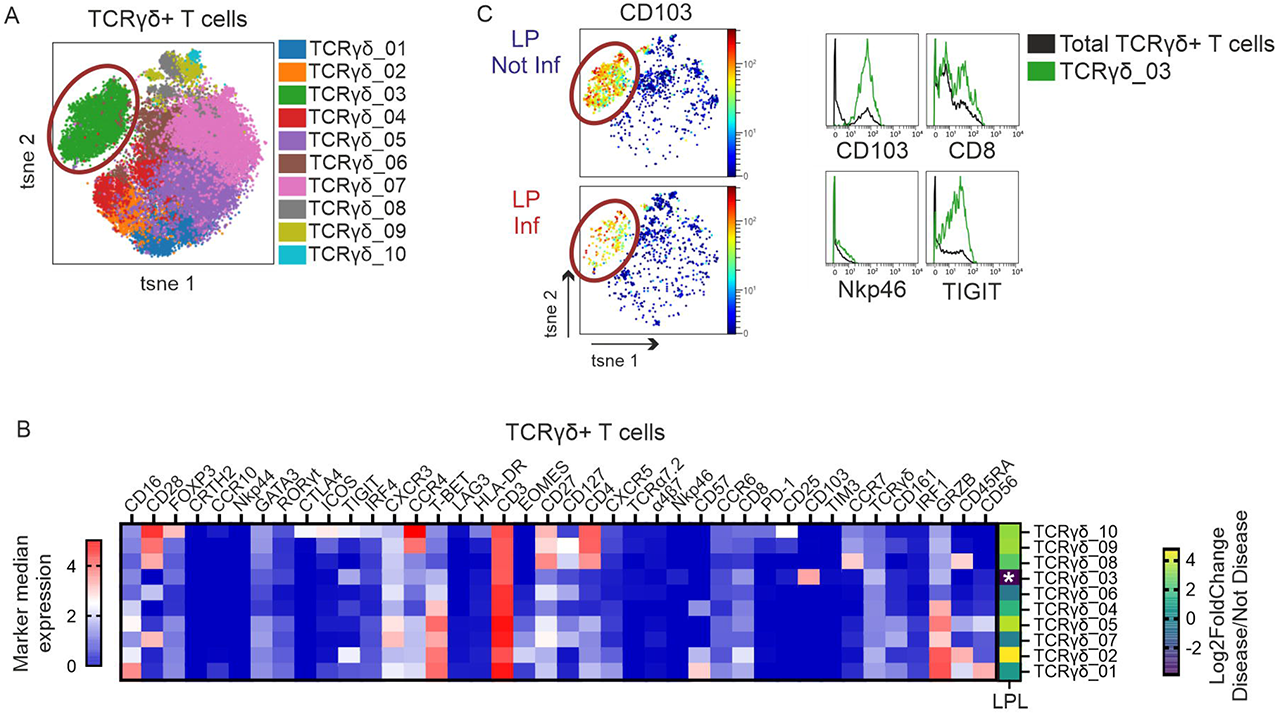
Loss of a tissue-protective CD103^+^NKp46^+^TCRγδ^+^ T-cell population in CD intestine. TCRγδ^+^ T cells were gated as shown in Figure S1A. (**A**) tsne, all samples merged, shows 10 TCRγδ^+^ T-cell clusters calculated with FlowSOM algorithm. Cluster TCRγδ_03 is highlighted. (**B**) Heatmap (calculated from all samples) of the 10 TCRγδ^+^ T-cell clusters (rows) shows median arcsinh-transformed signal intensities of 40 markers (columns) used for cluster analysis. The right part of the heatmap displays differences in abundance for each cluster (as % of CD3^+^ T cells) between LPL Inf and LPL Not Inf, as Log2FoldChange. Asterisks (*) indicate clusters significantly different between sample types. Statistic were calculated with Wilcoxon test and FDR correction for paired samples comparison. White asterisk: cluster significantly less abundant in LPL Inf vs Not Inf (FDR corrected p value < 0.05, Log2FoldChange < −0.6). (**F**) Left, CD103 expression intensity, shown in color continuous tsne plots, of LPL not inflamed vs. LPL inflamed samples. Right, histograms show median expression of main markers characterizing cluster TCRγδ_03.

### CD161^+^ and / or PD-1^+^ CD3^+^CD4^+^ T cells accumulate in inflamed intestinal areas, whereas innate-like CD3^+^CD4^+^CD16^+^CCR6^+^ T cells appear in the blood of CD patients

The distribution of CD3^+^CD4^+^ T cells (TCRγδ^+^ and MAIT cells excluded) was depicted in UMAP plots thereby revealing their phenotypical similarity. We overlaid CD4^+^ T cells of inflamed vs. not inflamed individual tissues (Figure 3A). Most phenotypes were detectable at all locations analyzed. However, CD103 expression was mostly restricted to intestinal CD4^+^ T cells, whereas EOMES and CD57 was almost exclusive to peripheral CD4^+^ T cells (Figure 3, A and B). We then further determined 30 CD4^+^ clusters by FlowSOM, of which CD4_18 was not further considered based on low CD3 expression (Supplemental Figure 4, A and B). 17 out of 29 CD3^+^CD4^+^ clusters were significantly altered in inflamed vs. not inflamed epithelia, 12 in LP, 7 in blood, mostly increased (all clusters analyzed as percentage of CD3^+^ T cells) (Supplemental Figure 5, A-C; Figure 3C). Consequently, the correlation landscape across clusters was distorted under conditions of inflammation (Supplemental Figure 6, A-C). Remarkably, all CD4^+^ Treg subpopulations were enriched in inflamed LP and epithelium, while the frequency of naive CD45RA^+^ Tregs, CD4_30, expanded in CD patients’ blood with duration of disease, although not generally with the age of patients (Figure 3D).

**Figure 3.**
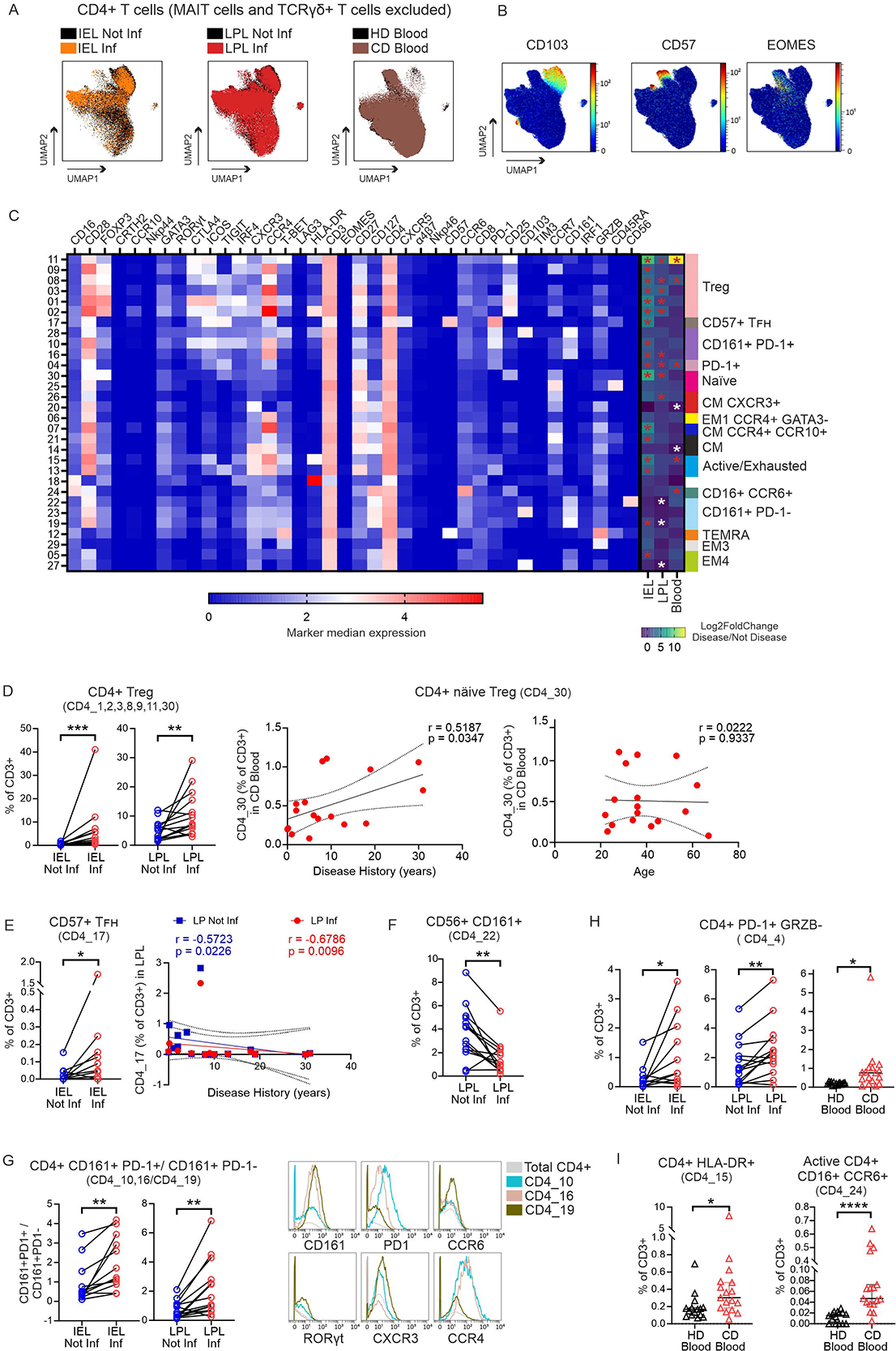
CD161^+^ and / or PD-1^+^CD3^+^CD4^+^ T cells accumulate in inflamed intestinal areas, whereas innate-like CD16^+^CCR6^+^ T cells appear in the blood of CD patients. (**A**) Tissue and disease-specific cell distribution of CD4^+^ T cells (MAIT cells and TCRγδ^+^ T cells excluded) on UMAP plots. Colors represent the different conditions as indicated. (**B**) Expression intensity, in UMAP plot, of tissue-specific markers. (**C**) Heatmap of FlowSOM identified 30 CD4^+^ T cells clusters (rows) shows median arcsinh-transformed signal intensities of 38 markers (columns) used for cluster analysis. Cluster annotations are given on the right. The right part of the heatmap further displays results of differential abundance analyses, i.e. Log2FoldChange for each cluster (as % of CD3^+^ T cells) between Inflamed vs Not inflamed (IEL and LPL) and CD blood against HD blood. Asterisks indicate clusters with significantly different abundance (white = lower; red = higher) between the conditions indicated. Wilcoxon test and FDR correction for paired intestinal samples comparison; Mann-Whitney test with FDR correction for unpaired blood samples. (FDR corrected p value < 0.05, Log2FoldChange < −0.6 and > 0.6). (**D**) Frequencies of CD4^+^ Tregs in CD intestine (left). Correlation of frequencies of näive Treg cells in CD blood with disease duration (center) and with patients’ age (right). (**E**) Frequencies of CD4^+^CD57^+^ T_FH_ cells in CD intestine (left) and their correlation with disease duration. (**D, E**) r and p values were determined based on Spearman’s correlation test, lines indicate linear regression. (**F**) Frequencies of CD4^+^CD56^+^CD161^+^ T cells. (**G**) Ratio between CD4^+^CD161^+^PD1^+^ and CD4^+^CD161^+^PD1^−^ T cells in CD intestine (left) and expression of selected markers that differentiate the identified CD4^+^CD161^+^PD1^+/–^ T cell clusters (right). (**H**) Frequencies of CD4^+^PD1^+^GRZB^−^ T cells in all tissues. (**I**) Frequencies of activated CD4^+^HLA-DR^+^ T cells (left) and active CD16^+^CCR6^+^ T cells (right) in blood. Each data point represents one patient. IEL, n=12; LPL, n=14; CD blood, n=17; HD blood, n=15. Statistics: two-tailed Wilcoxon test for paired intestinal samples and Mann-Whitney test between unpaired blood samples (*, p < 0.05; **, p < 0.01; ***, p<0.001; ****, p< 0.0001).

Cells in cluster CD4_17 expressed CXCR5, the homing receptor for germinal centers, as well as the T-follicular helper (T_FH_) cell-typical ICOS and PD-1. High levels of CD57 identified them as a subset of T_FH_-cells (26, 27). However, they also exhibiting CD45RA and could thus be defined as T_EMRA_. We found them enriched in inflamed intestinal epithelium, while they were lost in LP during the course of disease (Figure 3E). Moreover, we observed that IRF-1 expression, transmitting interferon signaling leading to Th1 or T_R_1 differentiation, but also innate-like phenotypes (28), was exclusive for activated CD4^+^ T cells, identified by either CD25 or HLA-DR expression, or for CD57^+^ T_EMRA_ (Supplemental Figure 7A).

CD161^+^CD56^+^CD103^lo^ NKT cells (CD4_22) were lessened in inflamed LP (Figure 3F). CD161 is a C-type lectin, known to be expressed by innate NK, innate-like MAIT, NKT, but also by subsets of CD4^+^ and CD8^+^ T cells with a Th17/Tc17 memory phenotype (29). Accordingly, six clusters were high in CD161, namely CD161^+^PD-1^+^ CD4_28, 10, 16 and CD161^+^ CD4_22, 23, 19, the three latter PD-1^−^ and CD127^hi^. The PD-1^lo/–^ CD4_19 expressed the highest levels of CCR6 and RORγt indicative of canonical Th17 cells (Figure 3G). Both, CD161^+^PD-1^+^ and CD161^+^PD-1^−^, tissue-resident CD103^+^ subpopulations (CD4_28, _23) did not change, while we detected more of the two other CD161^+^PD-1^+^ in inflamed epithelial and one also in LP. Thus, the ratio between CD4^+^CD161^+^CD103^−^ T cells shifted towards the ones expressing PD-1 due to inflammation in intestinal tissues (Figure 3G). In addition, PD-1^+^TIGIT^+^ICOS^+^GRZB^−^ CD4_04 were augmented at all sites upon inflammation (Figure 3H).

Finally, not only the extent of activated HLA-DR^+^ T cells, but specifically the innate-like cluster CD4_24 CD16^+^CCR6^+^ became apparent in CD patients’ blood, while being absent among HD PBMCs (Figure 3I).

### While T_RM_ cells disappear, highly activated T_EM_ and T_EMRA_ enrich in inflamed intestinal tissues and innate-like CD3^+^CD8^+^CD16^+^CCR6^+^ T cells emerge in the blood of CD patients

Next, we focused specifically on CD3^+^ CD8^+^ T cells. The UMAP plot depicted the overall division of CD8^+^ T cells in two large groups, either almost intestine-exclusive CD103^+^ or CD103^−^ CD8^+^ T cells (Figure 4, A and B). The overlaid cells from inflamed vs. not inflamed samples from each tissue showed small, but clear disease-related differences in cell distribution. FlowSOM identified 25 clusters, summarized in a heatmap (Supplemental Figure 8, A and B; Figure 4C). The calculated changes revealed a complex picture of enhanced and reduced subpopulation abundance due to inflammation (all clusters analyzed as percentage of CD3^+^ T cells) (Supplemental Figure 9, A-C), which again heavily disturbed the correlated ratio of cluster frequencies (Supplemental Figure 10, A-C). Like the CD4^+^ Treg subpopulations, CD8^+^FOXP3^+^ Tregs (30), CD8_24, was augmented in inflamed LP and epithelium (Figure 4D). Four, namely CD8_1, _2, _3 und _5, tissue-resident CD103^+^ subpopulations, considered to be the first line of defense (31), were less present upon inflammation, with a trend among IELs and statistical significance in inflamed LP (Figure 4E). Although in a varying extent, all four were additionally CD161^+^. A subpopulation with the highest CD161 level, but without the tissue-maintaining CD103 (32), expressed CD56 and could hardly be detected in blood of CD patients compared to HD (Figure 4F). CD8^+^CD161^+^ T cells are described as long-term memory cells with protective cytotoxic activity (33). On the contrary, the clusters with the three highly activated or even pre-exhausted CD8^+^ T cells (CD8_15, _18, _25) were significantly more abundant in inflamed epithelia and LP (Figure 4G). The frequency of the most activated ones according to HLA-DR expression levels, CD8_25, even accumulated in LP over the duration of CD.

**Figure 4.**
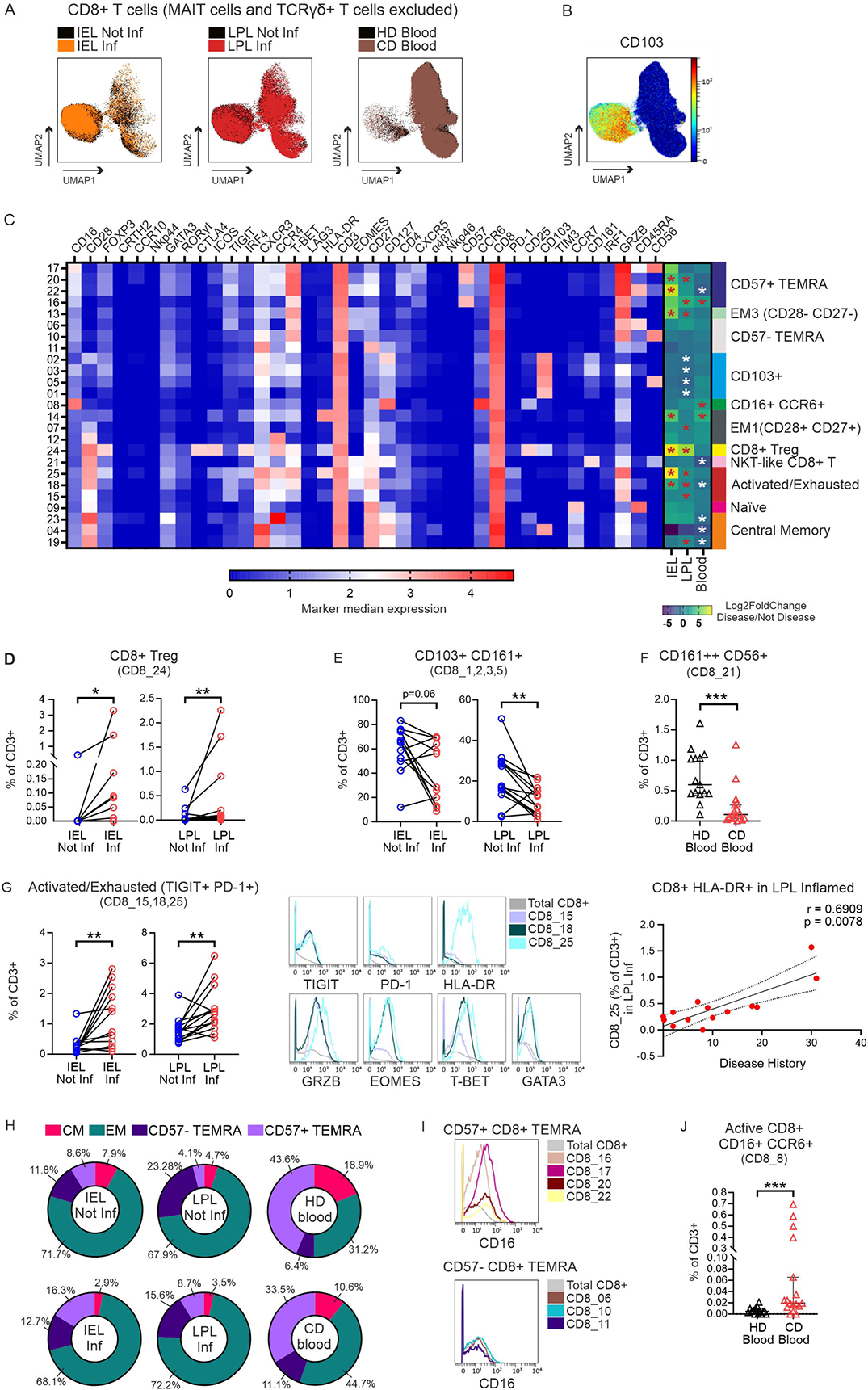
Accumulation of intestinal CD8^+^ T_EMRA_ and activated T_EM_ as well as peripheral innate-like T cells. (**A**) Tissue and disease-specific cell distribution of CD8^+^ T cells (MAIT cells and TCRγδ^+^ T cells excluded) on UMAP plots. Colors represent the different conditions as indicated. (**B**) Expression intensity, in UMAP plot, of CD103. (**C**) Heatmap of the 25 CD8^+^ T-cell clusters (rows) shows median arcsinh-transformed signal intensities of 38 markers (columns) used for cluster analysis. Heatmap calculated as in Fig.3C. (**D**) Frequencies of CD8^+^ Treg and (**E**) CD8^+^CD103^+^ CD161^+^ T cells in CD intestine. (**F**) Frequencies of CD8^+^ NKT-like T cells in blood. (**G**) Activated/exhausted CD8^+^TIGIT^+^PD1^+^ T cells in intestinal tissues (left); expression of key activation/exhaustion surface and intracellular markers (center) and correlation of HLA-DR^+^CD8^+^ T cells in inflamed LPL with disease history (in years). Spearman’s correlation test and linear regression (right). (**H**) Pie charts representing distributions of CD8^+^ T_CM_, T_EM_ and T_EMRA_ phenotypes in a tissue-dependent and disease-dependent manner. (**I**) Difference in CD16 expression between CD57^+^ and CD57^−^ T_EMRA_. (**J**) Frequencies of active CD16^+^CCR6^+^ T cells in HD and CD blood. Mann-Whitney test between unpaired blood samples. Each data point represents one patient (IEL, n=12; LPL, n=14; CD blood, n=17; HD blood, n=15). Statistics: two-tailed Wilcoxon test for paired intestinal samples and Mann-Whitney test between unpaired blood samples (*, p < 0.05; **, p < 0.01; ****, p< 0.0001).

Taking clusters of certain CD3^+^CD8^+^ memory subtypes together, we noticed that the frequency of T_CM_ reduced upon inflammation at all sites, leading to a significantly decreased ratio of T_CM_/T_EM_ in the blood of CD patients (Figure 4H, Supplemental Figure 9D). Besides the naïve-like CD8_09, we found seven CD45RA^+^ subpopulations defining CD3^+^CD8^+^ T_EMRA_ cells. CD8_16, _17, _20, _22 co-expressed the senescence marker CD57 (and some CXCR5), whereas CD8_6, _10, _11 did not. In inflamed epithelia and LP, CD57^+^ T_EMRA_ were relatively more abundant among CD8^+^ T cells and the ratio of CD57^+^ vs. CD57^−^ T_EMRA_ was significantly enhanced as compared to not inflamed tissues. In inflamed LP, also T_EM_ dominated over CD57^−^ T_EMRA_. As in CD4^+^ T cells, IRF-1 expression was exclusive for activated HLA-DR^+^CD8^+^ T cells and CD57^+^ T_EMRA_ (Supplemental Figure 7B). CD57^+^ T_EMRA_ cells expressed decent amounts of CD16 (Figure 4I). Cells with the highest level of CD16, however, could be identified as CD8^+^CD16^+^CCR6^+^ cells, making them a counterpart to the innate-like CD4^+^ T cells, correspondingly appearing in the blood of CD patients (Figure 4J). In sum, CD patients lost tissue-protective CD161^+^ cells, exhibited highly activated and / or pre-exhausted T_EM_ and CD57^+^ T_EMRA_ CD8^+^ cells in intestinal tissues, whereas an innate-like CD16^+^CCR6^+^ subpopulation appeared newly in the blood.

### Inflamed LP failed to keep a tissue-protective CD161^+^PD-1^+^ Treg subtype

Despite the more activated, exhausted and even aberrant phenotype of Tcon in CD patients, Tregs frequencies were clearly increased in inflamed epithelia and LP (Figure 5A). Only in CD patients’ blood, the pool of CD4^+^ Tregs were less, yet this one cluster of CD8^+^ Tregs was enriched here, too. To proof whether CD4^+^ Tregs disappear from the blood while migrating to the intestinal tissues, we assessed whether there was a negative correlation between Treg abundance in the inflamed tissue (LPL and IEL) and the blood across all CD patients, but could not find such a correlation (Supplemental Figure 11A).

**Figure 5.**
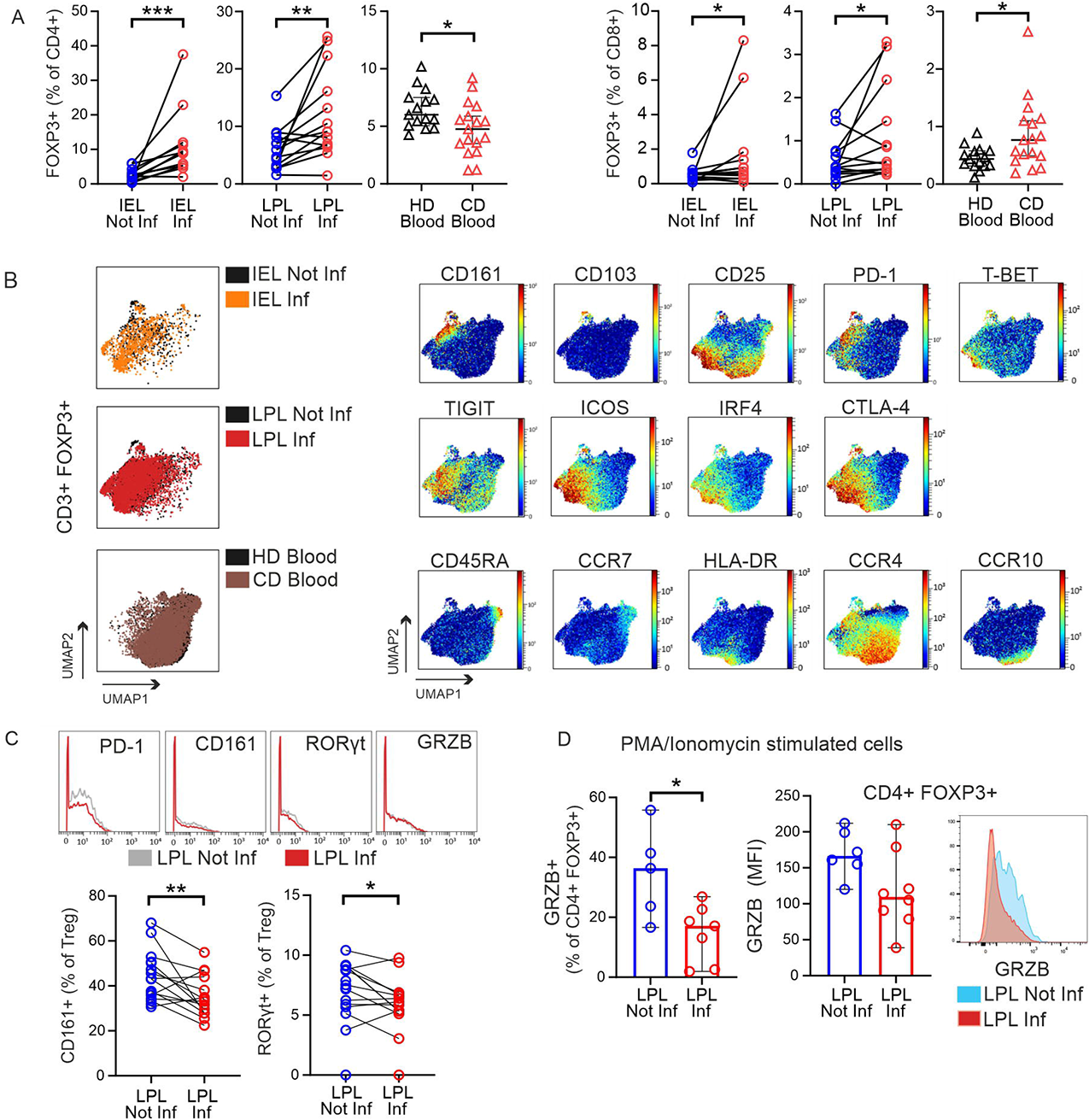
Expanded intestinal Tregs with apparent loss of protective function in CD patients. (**A**) Frequencies of FOXP3^+^CD4^+^ (left) and FOXP3^+^CD8^+^ (right) T cells were determined in mass cytometry data by manual gating in different tissues and disease statuses. Each data point represents one patient (IEL, n=12; LPL, n=14; CD blood, n=17; HD blood, n=15). (**B**) Left, tissue and disease-specific cell distribution in UMAP plots generated of CD3^+^FOXP3^+^ T cells manually gated (MAIT cells and TCRγδ^+^ T cells included). Colors represent the different tissues overlapped. Right, tissue-specific distribution of selected markers in UMAP of CD3^+^FOXP3^+^ from all concatenated samples (dark red, high expression; dark blue, no expression). (**C**) Overlay histograms of disease-specific selected markers on total CD3^+^ Tregs (top) and abundance of CD161^+^ and RORγt^+^ Treg in lamina propria (bottom). (**D**) Intestinal lymphocytes from a second cohort of CD patients were isolated, stimulated in vitro with PMA/Ionomycin in the presence of Brefeldin A and Monensin, and intracellular staining for GRZB was detected by flow cytometry. Frequencies of GRZB^+^CD4^+^ Tregs (left) and GRZB MFI (median fluorescent intensity) in Treg (center) in not inflamed (n=6) and inflamed (n=8) LPL cells. Each point represents one patient sample. Histogram shows intracellular GRZB staining in CD4^+^FOXP3^+^ T cells from one representative patients. Two-tailed Wilcoxon test was used for comparing paired intestinal samples and two-tailed Mann-Whitney test for unpaired blood samples (*p < 0.05, **p < 0.01, ***p<0.001).

To have an undisturbed view on Tregs only, we pre-gated CD3^+^FOXP3^+^ T cells and plotted them with UMAP (Supplemental Figure 11B). The FOXP3^+^ cells exposed different degrees of IL-2Rα/CD25 expression and no IL-7Rα/CD127, with the majority being CD4^+^ and only a small population CD8^+^. For further analyses, the contaminating CD127^+^ T cells were excluded (Supplemental Figure 11C). Of note, when we overlapped the Tregs in intestinal tissues and blood, the distribution of peripheral Tregs was different from that in the tissues (Figure 5B). Looking for tissue-specific features, we confirmed that tissue-healing CD161^+^(34) are exclusively present in the intestinal tissues. With PD-1, TIGIT, ICOS, IRF4 and CTLA-4, the intestinal Tregs exposed an effector signature (eTregs), supposingly being highly suppressive. On the other side, only in blood we found naive Tregs with CD45RA and CCR7 expression together with eTregs positive for HLA-DR and LP (or skin)-homing receptor CCR4 or CCR10 (Figure 5B).

When we now compared not inflamed vs. inflamed LP, we noticed that Tregs exhibited less PD-1 and CD161, i.e. had less tissue-remodelling capacity in the inflamed patches (Figure 5C). For CyTOF, cells had not been stimulated beforehand. Thus, we activated LPLs, collected from a second cohort of CD patients (Supplemental Table 2), by PMA/Ionomycin, analysed by fluorescence-based flow cytometry and now detected reduced GRZB expression (MFI) in LP CD4^+^FOXP3^+^ T cells and a lower frequency of GRZB^+^ LP Tregs from inflamed vs. not inflamed areas (Figure 5D). Less GRZB pointed to a reduced suppressive function. In sum, although Tregs were overall amplified in inflamed tissue areas, PD-1 and CD161 as well as GRZB expression was downregulated.

### An innate-like CD56^+^ Treg subtype with a pro-inflammatory phenotype enriched in inflamed intestinal tissue areas

Looking for Crohn’s disease-specific Tregs, we detected an untypical CD56^+^ subtype, appearing among CD3^+^CD4^+^FOXP3^+^ IELs and significantly increased among LPLs in inflamed regions (Figure 6, A and B). Since on the opposite, we had noticed that CD4^+^CD56^+^ NKT cells were reduced in the same area (Figure 3C, CD4_22), we calculated the ratio. Clearly, CD56^+^ Tregs dominated over all CD4^+^CD56^+^ non-Tregs in inflamed epithelia and LP (Figure 6B). To ensure the presence of such a small and unusual Treg subtype, we re-evaluated former CyTOF data of IBD patients shared in the Cytobank repository (19). Reassuringly, CD4^+^FOXP3^+^CD56^+^ cells could be found in biopsies of LP of CD and UC patients (Supplemental Figure 12). Interestingly, there were even more such Tregs in inactive than active disease, whereas the ratio of CD4^+^FOXP3^+^CD56^+^ Tregs to CD4^+^FOXP3^−^CD56^+^ Tcon seemed to be highest in active IBD. Now, we isolated LP lymphocytes from the second cohort of CD patients, stimulated them in vitro and demonstrated per conventional flow cytometry not only the presence of such CD56^+^ Tregs, but their accumulation upon inflammation (Figure 6C). To reveal, whether CD56^+^ Tregs are more related to classical Tregs or to CD56^+^ Tcon, we compared the overall expression of all the acquired markers in a heatmap for those three populations (Figure 6D). Obviously, CD56^+^ Tregs are a subtype of Tregs and not a FOXP3-upregulating CD56^+^ Tcon. For functional analyses, we performed intracellular staining of in vitro stimulated T cells derived from inflamed parts of LP. CD56^+^FOXP3^−^ secreted the most IFN-γ and hardly any IL-10, whereas classical CD56^−^FOXP3^+^ Tregs showed the opposite cytokine profile (Figure 6E). While the percentage of IL-10^+^CD56^+^FOXP3^+^ Tregs equalled that of the IL-10^+^CD56^−^FOXP3^+^ Tregs, the CD56-positive Tregs also produced IFN-γ. Detecting GRZB and CD107a in parallel, CD56^+^FOXP3^+^ Tregs seemed to have a high cytolytic activity according to CD107a expression (Figure 6F). Taken together, these inflammation-enhanced innate-like Tregs exerted a pro-inflammatory phenotype.

**Figure 6.**
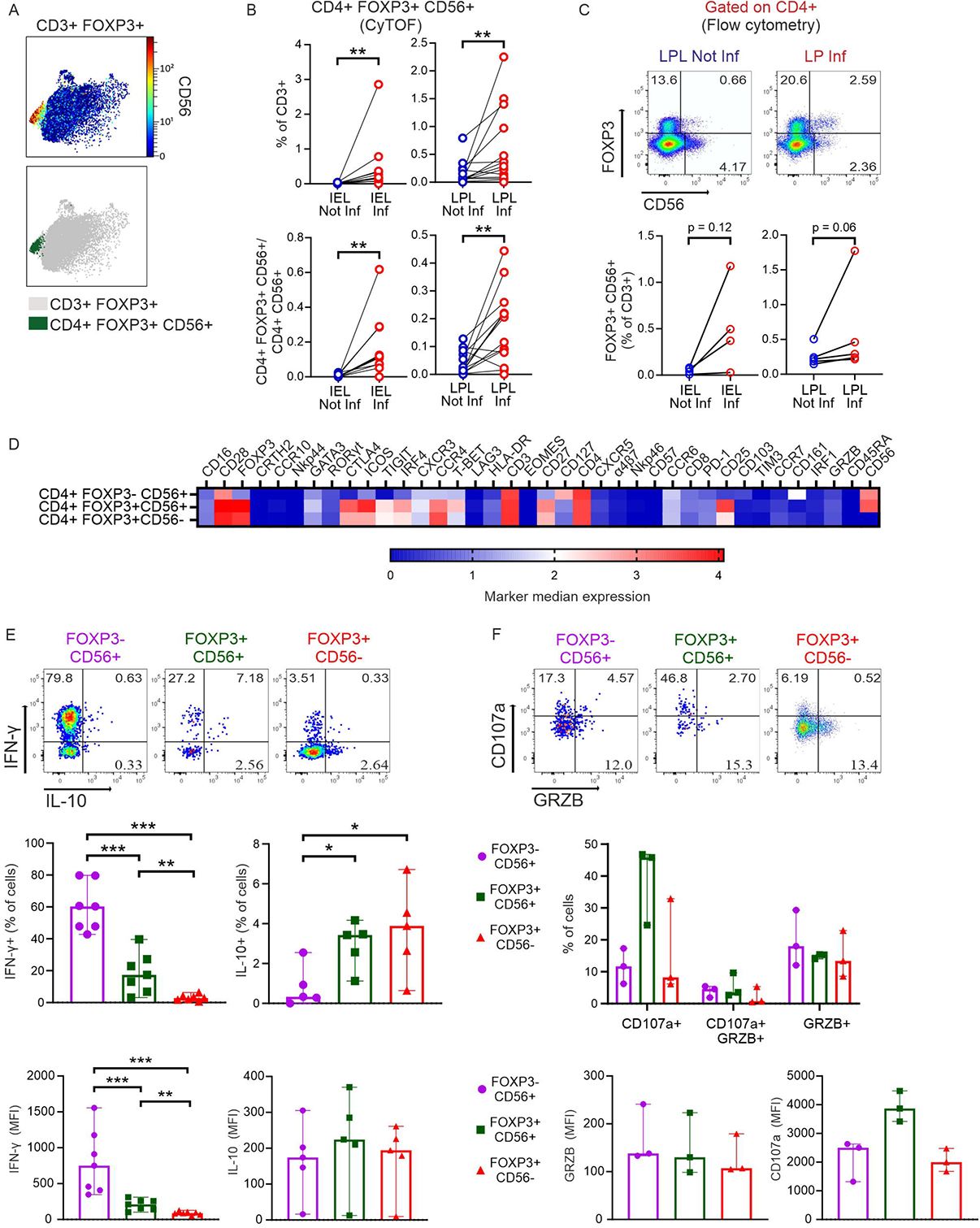
An innate-like CD56^+^ Treg subtype with a pro-inflammatory phenotype is enriched in inflamed intestine. (**A**) Top, CD56 expression by CD3^+^FOXP3^+^ T cells. Data from all concatenated samples are shown. Bottom, the same plot highlights CD4^+^FOXP3^+^CD56^+^ T cells, manually gated. (**B**) Frequencies (from MC data) of CD4^+^FOXP3^+^CD56^+^ T cells (top) and ratio of CD4^+^FOXP3^+^CD56^+^/CD4^+^CD56^+^ T cells (bottom) in intestinal samples. Two-tailed Wilcoxon test. (**C**) Identification of the CD4^+^FOXP3^+^ T cells expressing CD56 in a second group of CD patients by flow cytometry. Top, representative dot plots depicting data of LPL cells from one donor are shown as example. Bottom, frequencies of CD4^+^FOXP3^+^CD56^+^ in intestinal samples. Two-tailed Wilcoxon test. (**D**) Heatmap (from all intestinal samples) showing median arcsinh-transformed signal intensities of 38 markers (columns) among the innate-like FOXP3^−^CD56^+^ T cells, the newly identified FOXP3^+^CD56^+^ T cells and the classical FOXP3^+^CD56^−^ Tregs (rows) from MC data. (**E**) Intestinal lymphocytes from a second cohort of CD patients were isolated, stimulated in vitro with PMA/Ionomycin in the presence of Brefeldin A/Monensin and intracellular staining was used to detect the induction of the indicated cytokines by flow cytometry. Top, representative analysis of an LPL Inf sample of IL-10 and IFN-γ expression by the three populations in (D). Numbers indicate frequencies. Frequencies of IFN-γ^+^ and IL-10^+^ cells (center) and MFI of IFN-γ and IL-10 (bottom) within each population in inflamed LPLs. (IFN-γ, n=7; IL-10, n=5). (**F**) Top, representative analysis of an LPL Inf sample of CD107a and GRZB expression by the three populations in (D) (anti-CD107a added during stimulation). Numbers indicate frequencies. Frequencies of CD107a^+^ and GRZB^+^ cells (center) and MFI of CD107a and GRZB (bottom) within each population in LPL Inf (n=3). Two-tailed Mann-Whitney test (*p < 0.05, **p < 0.01, ***p<0.001).

### A TIGIT^−^HLA-DR^+^ Treg subtype appeared in inflamed intestinal tissue areas, expressed CD16 and had a pro-inflammatory phenotype

Another small, but seemingly interesting Treg subtype was TIGIT^−^HLA-DR^+^ (Figure 7A), the only group of intestinal Tregs negative for TIGIT. Again, it emerged upon inflammation within the CD3^+^ T-cell populations and correspondingly within the CD4^+^FOXP3^+^ subsets in epithelia and LP population (Figure 7B). Like before, also this Treg subpopulation was assessed by conventional flow cytometry. Within the gate of CD4^+^FOXP3^+^ cells, we detected TIGIT^+^, HLA-DR^+^, double-negative, and double-positive Tregs, subsequently calculating their abundance within CD3^+^ T cells (Figure 7C). Portraying the 40 marker expressions from the four populations in a heatmap revealed that TIGIT^−^HLA-DR^+^FOXP3^+^ cells not only exhibited the highest levels of T-BET, but the most pronounced CD16, once more representing an innate-like phenotype (Figure 7D). Accordingly, TIGIT^−^HLA-DR^+^ Tregs produced the highest amount of pro-inflammatory IFN-γ when comparing those four TIGIT and HLA-DR positive and negative Treg subgroups (Figure 7E). In sum, innate-like TIGIT^−^HLA-DR^+^CD16^+^ Tregs appeared in inflamed intestinal areas of CD patients exerting an elevated pro-inflammatory phenotype.

**Figure 7.**
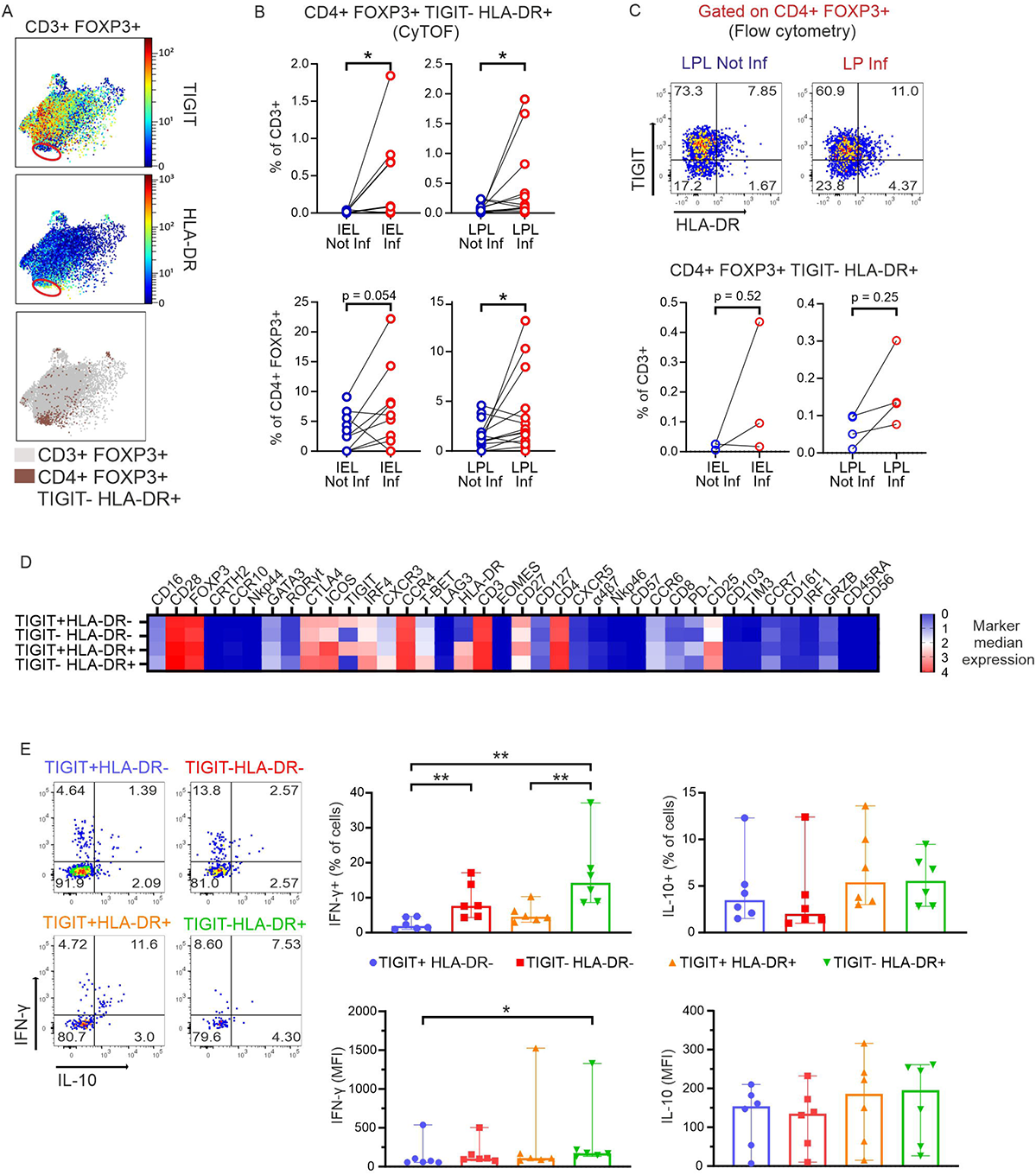
TIGIT^−^HLA-DR^+^ Tregs with a pro-inflammatory phenotype appear in inflamed CD intestine. (**A**) Top, TIGIT and HLA-DR expression by CD3^+^FOXP3^+^ T cells. Data from all concatenated samples are shown. Bottom: the same plot highlights HLA-DR^+^TIGIT^−^ Tregs. (**B**) Frequencies (from MC data) of CD4^+^FOXP3^+^HLA-DR^+^TIGIT^−^ T cells in intestinal samples within total CD3^+^ T cells (top) and within CD4^+^FOXP3^+^ T cells (bottom). Two-tailed Wilcoxon test. (**C**) Identification of the CD4^+^FOXP3^+^HLA-DR^+^TIGIT^−^ T-cell population in a second group of CD patients by flow cytometry. Top, representative dot plots depicting data of LPL cells from one donor are shown as example. Bottom, frequencies of CD4^+^FOXP3^+^HLA-DR^+^TIGIT^−^ T cells identified by flow cytometry in intestinal samples. Two-tailed Wilcoxon test. (**D**) Heatmap (from all intestinal samples) compares median arcsinh-transformed signal intensities of 38 markers (columns) among 4 Treg populations distinguished according to TIGIT and HLA-DR expression (rows) from MC data. (**E**) Intestinal lymphocytes from a second cohort of CD patients were isolated, stimulated in vitro with PMA/Ionomycin (plus Brefeldin A/Monensin) and intracellular cytokine staining was detected by flow cytometry. Left representative analysis of an LPL Inf sample of IL-10 and IFN-γ expression by the populations in (D). Numbers indicate frequencies. Right: frequencies of IFN-γ^+^ and IL-10^+^ cells and MFI of IFN-γ and IL-10 within each population in LPL Inf (n=6). Two-tailed Mann-Whitney test (*p < 0.05, **p < 0.01, ***p<0.001).

### Innate-like CD16^+^CCR6^+^ T cells can be induced by CD patient’s sera

So far, we got the impression that lately differentiated adaptive T cells can acquire expression of receptors known from innate immune cells and that this might be provoked by the pro-inflammatory situation of chronic CD. Accordingly, both CD4^+^ and CD8^+^, robustly positive for CD16, CCR6, CD127 and CD25, could only be detected among CD patients’ PBMCs, but not in HD blood (Figure 8A, Figures 3I and 4J), while present in both inflamed and not inflamed intestinal samples from CD patients (Supplemental Figure 13). To test whether CD patients’ blood contains inducing factors, we incubated CD3^+^ T cells from HD with plasma from CD – and here included UC – patients and HD in vitro. CD3^+^ T cells were stimulated by anti-CD3/CD28 in the presence of IL-2 and the differently derived plasma. Indeed, compared to plasma from HD, IBD patients’ plasma had induced CD16 and CCR6 on CD4^+^ T cells already after three days and a little delayed on CD8^+^ T cells after seven days (Figure 8B). These data indicated that soluble factors are responsible for promoting this aberrant innate-like phenotype of adaptive T cells. In line, the second cohort of IBD patients exposed significantly augmented frequencies of CD4^+^CD16^+^CCR6^+^ T cells among PBMCs of CD, but not UC patients (Figure 8C). Together, this implicated that the altered milieu in IBD patients further provokes changes in the composition of intestinal T-cell subtypes for which peripheral CD16^+^CCR6^+^ innate-like T cells can serve as a marker.

**Figure 8.**
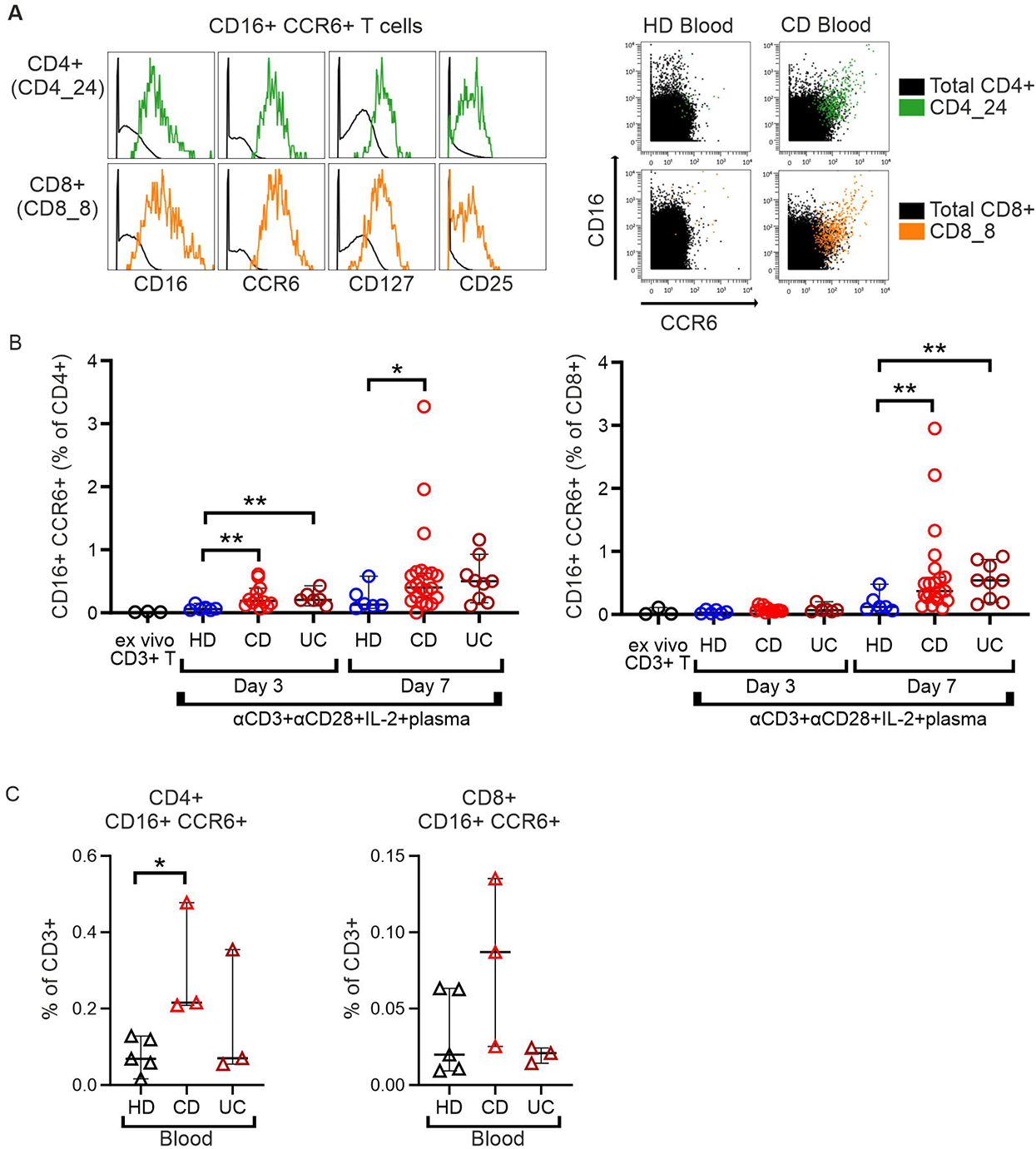
Innate-like CD16^+^CCR6^+^ T cells can be induced by CD patients’ sera. (**A**) Left, overlay histograms demonstrating expression of CD16, CCR6, CD127 and CD25 on CD4_24 and CD8_8 T-cell clusters. Right, overlaid CD and HD blood samples in two-dimension dot plots show the expression of CD16 and CCR6 of the identified cell clusters. (**B**) Differentiation of CD16^+^CCR6^+^ (CD4^+^ or CD8^+^) T cells in healthy donors’ isolated CD3^+^ T cells cultured, for 3 and 7 days, with 10 % of HD, CD (n=7) or UC (n=3) patients’ blood plasma. Experiments on 3 healthy donor CD3^+^ T cells, each cultured with the same CD (n=7) or UC (n=3) patients’ blood plasma, are pooled. Two-tailed Mann-Whitney test. (**C**) Frequencies of CD16^+^CCR6^+^ (CD4^+^ or CD8^+^) T cells in freshly isolated PBMCs from HD (n=5), CD (n=3) and UC (n=3) donors. Two-tailed Mann-Whitney test. (*, p < 0.05; **, p < 0.01).

## Discussion

The primary site of inflammation in IBD is the intestine, where the mucosal barrier should be maintained to control pathogens while being tolerant towards commensals. Integrity loss of this single layer of epithelial cells leads to entry of luminal content into LP and a subsequent inflammatory immune response by tissue-resident and immigrant immune cells, decisively driven by T cells (22, 23). Therefore, we analyzed CD3^+^ T cells within IELs and LPLs as well as PBMCs by mass cytometry / CyTOF. Disease vs. non-disease was evaluated by blood samples of CD patients in comparison to matched healthy donors, whereas intestinal sampling elicited T-cell compositions in actually inflamed LP and epithelia in comparison to macroscopically intact, but still patients’ tissue. In agreement with prolonged antigen exposure, various NK-cell receptors were upregulated on CD8^+^ T cells as described for other clinical circumstances like chronic virus infection, autoimmune diseases or tumors (35, 36). This implies eventual activation by cytokines, in the context or even absence of TCR signaling as well as innate functions of such CD8^+^ T cells at inflamed sites. In CD, also some CD4^+^ Tcon and Tregs exhibited innateness.

Overall and like others (20), we observed that CD8^+^ T cells were significantly decreased, while CD4^+^ T cells were increased within the CD3^+^ T-cell population in the inflamed epithelium. However, upon inflammation, innate CD3^−^ T cells outcompeted the overall number of CD3^+^ T cells already indicating a dominant innate, i.e. less specific immune reaction which coincides with a general loss of barrier protection by the adaptive immune system in CD. In line, inflammation influenced the cell composition in the LP where a CD103^+^ TCRγδ^+^ T_RM_ cell subtype diminished. Being positive for NKp46, CD8, CD103 and CD56, it resembled the largest subset of γδ T cells among intestinal IELs, described as a protective Vδ1 subpopulation in humans (25). We do not know the identity of the TCR chains, but assume them to be T_RM_-typical Vδ1^+^. The additional high TIGIT expression represented an activated phenotype.

Along with CD103^+^ γδ T cells, several subpopulations of CD103^+^CD8^+^CD161^+^ T_RM_ cells were lost in inflamed epithelia and LP. CD8^+^ T_RM_ cells support tissue homeostasis by exerting protective cytotoxicity. CD161 expression additionally provides gut-specific homing properties to T cells (37). CD8^+^CD161^+^ are long-lived memory cells, highly cytotoxic or – when CD161^hi^CCR6^+^ – IL-17-producing Tc17 cells (33). High levels of the chemokine receptor CCR6 decide over precise localization inside the intestinal microenvironment. There, they are pro-inflammatory and destructive, whereas a mislocalization of CCR6^−^ Th17 provokes a Th1 dominance and less Tregs (37, 38). We found an enrichment among both inflamed IELs and LPLs, interestingly especially of PD-1-expressing CD161^+^CCR6^+^ Th17. While IL-17A overexpressed in the stenotic intestine is already pro-fibrotic (39), PD-1^+^ Th17 were described to additionally secret high levels of TGF-β, which together with IL-17A promotes lung fibrosis (40). Intestinal fibrosis is a critical determinant of a CD patient’s prognosis and the high frequency of CD161^+^CCR6^+^PD-1^+^ Th17 in inflamed tissues correlated well with the necessity of surgical intervention in our study. The authors of the lung study assumed that those PD-1^+^ Th17 cells are converted Tregs (40).

Conformingly, Tregs have been described to transdifferentiate to T_FH_ cells, something exclusive for the gut-associated lymphoid tissue (41). The PD-1^hi^ICOS^+^ T_FH_ subset we observed diminished in the LP, where T_FH_ cells would be expected, over disease duration, but enriched in inflamed epithelia. High levels of CD57 hinted to a weak cytotoxic phenotype (26) or just terminally differentiated T_FH_ cells promoting plasma cell differentiation (27). In line with the latter, they were CD45RA^+^, i.e. T_EMRA_, and therefore probably less capable of a normal germinal center response. In parallel, the frequency of PD-1^+^TIGIT^+^ICOS^+^CXCR5^−^ cells correlated positively with inflammation at all three sites. Those could be functionally impaired, but proliferating CD4^+^ cells (42) or autoimmune disease-related CXCR5^−^ T-peripheral helper (T_PH_) cells. There have been hints for the existence of T_PH_ cells in CD patients already (21). Although T_PH_ cells exert T_FH_-like functions in inflamed tissues – mostly outside ectopic lymphoid aggregates – this leads to autoantibody production (43). In fact, mucosal barrier damage and bacteremia provoke anti-commensal antibody generation, which might recognize tissue antigens due to molecular mimicry (44). Thus, it appears that in CD malfunctioning T_FH_ and autoimmune disease-driving T_PH_ cells dominate as B-cell helpers.

Even if some Tregs converted to PD-1^hi^ Th17 and / or PD-1^hi^ T_FH_ cells, FOXP3^+^ Tregs were still abundantly present in CD patients and enriched in inflamed tissues. Although early studies had described defects in the number and distribution of Tregs in IBD, this was followed by the observation of expanded intestinal Tregs in CD patients (5, 45, 46). On top, we noticed a strong increase of naive Tregs in CD in comparison to HD blood. Together, this might reflect the attempt of the immune system to control the exuberant inflammation with the help of Tregs, something already in vain due to the described resistance of CD patients’ Tcon to Treg-mediated suppression (47). In addition, we found the protective Treg phenotype expressing PD-1, CD161 and GRZB to be underrepresented in inflamed LP. CD161^+^ Tregs had been described as partly pro-inflammatory (48), which could then be clarified as epithelial barrier-healing for the gut exerted by Th17-type cytokines (34).

Generally, Tregs can differentiate into all kinds of Th-like eTregs when driven by the respective Th-polarizing cytokines, then co-expressing the lineage-determining key transcription factors and chemokine receptors, while FOXP3 represses cytokine genes (49). Under pro-inflammatory conditions, however, Tregs might diminish the expression of their decisive transcription factor FOXP3 due to epigenetic changes and acquire features of pro-inflammatory cells (50, 51), also relevant for CD (52). In line, when we analyzed two non-classical Treg subtypes, both enriched in inflamed vs. not inflamed epithelia and LP, their dysfunctional IFN-γ^+^ Th1-type became obvious. One Treg subtype stained positive for CD56 (NCAM1). CD56^+^Foxp3^+^ Tregs were reported in the context of other diseases (53–55). All studies confirmed suppressiveness as well as induction by and / or production of TGF-β rather favoring the idea of FOXP3 induction in CD56^+^ Tcon. However, given the observed similarity of CD56^+^FOXP3^+^ Tregs in CD with classical Tregs, we assume an upregulation of CD56 on Tregs by activation and IL-12/23 or IL-15 as known for Tcon (56). Accordingly, analogous IFN-γ production is described for CD56^+^ Tcon and NK cells, which exert effector functions by Th1-type cytokines and cytotoxicity. Of note, CD56^+^ immune cells are able to form strong immune synapses with each other through homophilic CD56 binding, which could hint to an innate-like, TCR-peptide MHC-independent cell-cell contact of those CD56^+^ Tregs.

Likewise, the other discovered inflammation-induced and IFN-γ-secreting Treg subtype exposed an unusual phenotype. Although being an activated eTreg, i.e. HLA-DR^+^, it was TIGIT-negative. The checkpoint receptor TIGIT directs the suppressive function of eTregs (57), while a relative loss of TIGIT^+^ eTregs has been described for chronic diseases (58). Interestingly, the inflammation-associated TIGIT^−^HLA-DR^+^ Tregs exhibited some CD16 expression, again an innate-like receptor implying a differential cellular crosstalk.

CD16 is the low-affinity FcγRIII mediating antibody-dependent cellular cytotoxicity (ADCC) (59). It associates with ITAM (immunoreceptor tyrosine-based activation motif)-containing FcRγ and / or CD3ζ for signal transduction (60). Besides those aberrant TIGIT^−^CD16^+^ eTregs, we noticed CD16^+^CCR6^+^ Tcon, either CD4^+^ or CD8^+^, appearing among PBMCs in CD patients and present in inflamed as well as not inflamed intestinal tissue areas of CD patients. Highly cytotoxic CD16^+^CD8^+^ T_EMRA_were identified in hepatitis C virus-infected patients, kidney transplant recipients at high risk of graft failure as well as in smokers (61–63), in conjunction with our results correlating smoking-mediated senescence-associated immune dysregulation with IBD. However, a tiny fraction of CD4^+^ and CD8^+^CD16^+^ T_EM_ cells was also observed in HD (60, 64). CD8^+^CD16^+^ T_EM_ are capable of mediating ADCC immediately ex vivo (64), whereas CD16 expression on activated peripheral CD4^+^ T cells serves as co-stimulus for high IFN-γ or IL-17 expression. Here, a non-lytic complement complex might form a synapse facilitating the deposition of immune complexes and subsequent signaling (60). Remarkably, CD16^+^CD4^+^ and CD16^+^CD8^+^ T_EM_ cells (both with a cytotoxic phenotype and not T_EMRA_) are strongly amplified in COVID-19 patients, their numbers correlating with severity of disease (65). The authors describe a complement-dependent – likely involving the complement receptors C3a and C3b – induction of CD16, upon which those adaptive T cells exert immune complex-mediated, TCR-independent cytotoxicity (65). In accordance with a hyper-activated complement system in chronic inflammatory diseases including CD (66, 67), we were able to induce CD16 expression on HD-derived T cells by CD patients’ plasma. Analogous to the CD16^+^CCR6^+^CD4^+^ and CD16^+^CCR6^+^CD8^+^ T_EM_ cells, which we found in CD patients, the ones in COVID-19 patients are highly CCR6 or CXCR3-positive, there leading them to the lung, while we anticipate a preferential homing to the intestines during IBD. Because of barrier breakdown and leakage of luminal content, (complement-opsonized) immune complexes are abundantly present in CD (44), now becoming a device for TCR-independent, innate-like and possibly tissue-destructive actions of formerly adaptive T cells. Depending on the FcγRIII-bound (auto-) antibodies (44), possibly enabled by T_PH_ and CD45RA^+^CD57^+^ T_FH_ cells, CD16^+^CCR6^+^ T cell-mediated cytotoxicity will lead to various tissue destructions, which might even provoke endothelial dysfunction like in COVID-19 patients (65, 68), foreshadowing extra-intestinal manifestations of CD.

Taking the mentioned reports together while including ours, it appears that the transformation of adaptive T cells into innate-like NK cell-resembling cells is one underlying mechanism for immunopathology in chronic inflammatory diseases like CD.

## Material and Methods

### Study with cells from intestinal and blood samples from human subjects

A total of 15 healthy individuals and 25 CD patients (17 in the first, 8 in the second cohort; Supplemental Table 1 and 2) were included in the study. From CD patients undergoing ileocecal resection surgery, intestinal samples from inflamed and not inflamed regions were resected and directly transferred into PBS and kept on ice until further processing. Blood from CD patients and HD were collected in NH4-Heparin tubes (Sarstedt). The median age of healthy donors is 37 (22–67) years and the median age of CD patients is 39 (23–62) years. Both groups contain males (59%) and females (41%). All the patients had similar treatment history, with corticosteroids and anti-TNF-α monoclonal antibodies; only four patients (2 in Cohort 1 and 2 in Cohort 2) were treated with anti-α4β7 monoclonal antibodies (Vedolizumab). Details are provided in Supplemental Tables 1 and 2.

### Human intestine dissociation and cell isolation

IEL and LPL from surgically resected terminal ileum samples (inflamed and not inflamed) were isolated as previously described (69). In brief, freshly resected tissues were kept in PBS on ice and transferred to the lab in less than one hour. Fat tissue was carefully removed and the intestinal samples were minced in smaller pieces using a scalpel. Epithelial cells were isolated by two washing steps in HBSS, 5 % FCS, 5 nM EDTA and 2 mM DTT by shaking at 37°C, 20 min, 150 RPM. Successively, remaining LP was minced and enzymatically digested, in two steps, with 0.5 mg/ml Collagenase D (Roche) and 0.1 mg/ml DNase I (Roche) by shaking at 37°C, 40 min, 250 RPM. Finally, both epithelial and LP cells were fractionated through a 40 %/80 % gradient centrifugation of Percoll® (Sigma-Aldrich) for purification of IEL and LPL. PBMC were isolated from whole blood by density gradient centrifugation over Ficoll-Paque (Ficoll® Paque Plus, Cytiva) according to manufacturer’s manual. For mass cytometry, isolated cells were cryopreserved in freezing medium (90 % FCS mixed with 10 % DMSO) in liquid nitrogen. For flow cytometry analysis, cells were rested overnight, at high density (≈10^7^ cells/ml) in TexMACS™ Medium (Miltenyi Biotec) with 10 % FCS, 0.1 mM β-mercaptoethanol (Gibco), 1 mM sodium pyruvate, 0.1 mM nonessential amino acids, 100 U/ml penicillin, 100 µg/ml streptomycin, and 10 mM Hepes (Invitrogen), in a humidified incubator maintained at 37°C with 5 % carbon dioxide atmosphere.

### Antibodies and reagents for mass cytometry

Antibodies (Supplemental Table 3) were obtained pre-conjugated to metal isotopes (Fluidigm) or in purified form from Biolegend, Miltenyi Biotec, BD Biosciences, Thermo Fisher Scientific, R&D Systems, Sanquin, or were produced in-house (DRFZ), and conjugated in-house using the MaxPar X8 kit (Fluidigm) following the manufacturer’s instructions. Metal isotopes/elements not available from Fluidigm were purchased from Trace Sciences or Sigma-Aldrich. Isotopically enriched cisplatin was purchased from or kindly provided by Fluidigm. Palladium and cisplatin antibody conjugations were performed in-house as described before (70, 71). Fluorochrome-conjugated antibodies were purchased from Miltenyi Biotec and Cell Signaling Technologies. Fc-blocking reagent was purchased from Miltenyi Biotec. All antibody master-mixes were prepared once and aliquots were cryopreserved at −80°C as described before (72). Prior to use, thawed antibody master-mixes were centrifuged (4°C, 15000 xg, 10 min) to pellet potential aggregates. mDOTA-103Rh, used for discrimination of dead cells, was prepared from DOTA-maleimide (Macrocyclics, Dallas, TX) and rhodium chloride (Sigma-Aldrich) as previously described (73) and stored at 4°C.

### Mass cytometry staining and data acquisition

IELs (inflamed and not inflamed), LPLs (inflamed and not inflamed) and PBMC from 17 CD patients (first cohort) and PBMCs from HD were acquired in 4 individual batches. One PBMC sample was chosen as anchor control and was included in each batch. The mass cytometry staining was performed as previously described (24). In brief, cryopreserved cells from 20 samples were thawed in pre-warmed (37 °C) complete RPMI (Sigma-Aldrich; RPMI 1640, 10 % FCS, glutamine, penicillin, and streptomycin), supplemented with 2.5 U/ml Benzonase HC (Millipore), and re-suspended in CyTOF staining medium (1x PBS (prepared from 10x PBS (Rockland Immunochemicals), 0.5 % (w/v) BSA (PAN-Biotech), and 0.02 % sodium azide (Sigma-Aldrich)) containing 0.5 U/ml Benzonase HC (CSM+B). Cells samples were barcoded by incubation with premixed combinations of 2 out of seven metal-conjugated β-2-microglobulin (B2M) antibodies (89Y-,104Pd-,108Pd-,110Pd-,195Pt-,196Pt-, and 198Pt) (70, 74) supplemented with 1 μl Fc-blocking solution (Miltenyi Biotec), and CD19-, CD20-, CD36-, and CD123-PE conjugates. Following the barcode staining, the cells were washed and all samples were pooled for the subsequent staining steps. Pooled cells were incubated in cell-surface antibody cocktail (Supplemental Table 3) for 30 min at 4° C and dead cells labelled by incubation with 103Rh-mDOTA at room temperature (RT) for further 5 min. Afterwards, cells were washed with CSM and permeabilized using FoxP3 Staining Buffer Set (Miltenyi Biotec) according to manufacturer’s instructions. Next, incubation (30 min at RT) with a cocktail of antibodies directed against intracellular markers (Supplemental Table 3) was performed, followed by a washing step with Permeabilization Buffer (Miltenyi Biotec) and 172Yb-labeled anti-Cy5 antibody staining. Finally, cells were washed and incubated in 2 % paraformaldehyde (Electron Microscopy Sciences) at 4° C, overnight. On the next day, cells were washed once in CSM and then incubated for 25 min, at RT in iridium-based DNA intercalator solution (Fluidigm) (diluted 1/500 v/v from 0.125 mM in PBS). Afterwards, cells were counted on a MACSQuantflow cytometer (Miltenyi Biotec), washed and resuspended in an appropriate volume of water to obtain a maximum of 7.5 × 10^5^ cells/ml and finally supplemented with EQ Four Element beads (1/10, v/v) (Fluidigm). Next, cell acquisition was performed at an injection rate of 30 μl/min on a Helios mass cytometer (Fluidigm), tuned, cleaned and operated according to the manufacturer’s advice, with appropriate washing and tuning solutions (Fluidigm), on the day of the measurement. The mass cytometer was run in dual calibration mode, with noise reduction turned on and event length thresholds set to 10 and 75.

### Mass cytometry data analysis

Raw mass cytometry data were converted in Flow Cytometry Standard (FCS) 3.0 files during acquisition. Data were normalized for time-dependent signal drift based on EQ four element calibration beads, using the CyTOF acquisition software (v7.0)(Fluidigm). Next, spillover compensation was performed, using the CATALYST package (75) in R (R version 3.6.1; R Studio 1.2.5). The applied spillover matrix was generated previously, by Budzinski et al. (76), with BD Compensation beads and a set of antibody CyTOF conjugates based on the same metal stocks.

Data were manually de-barcoded using FlowJo (version 10.7.1) (FlowJo LLC, Ashland, OR) as described before (70). The data of individual cell samples were then imported into OMIQ.ai (Santa Clara, CA), where single, live, CD45^+^ cells were gated according to 103Rh-mDOTA for dead cell exclusion, DNA and event length parameters, and CD45 expression (Supplemental Figure 1A) for further analysis. Samples with rather few cell events were excluded from the subsequent analysis (final analysis was carried out on n=16 IEL Not Inf; n=12 IEL Inf; n=16 LPL Not Inf; n=14 LPL Inf; n=16 CD blood; n=15 HD blood). Data visualizations, t-SNE plots (t-SNE coordinates calculated with opt-SNE (77), FlowSOM clustering (78) and further manual gating was performed in OMIQ. Dot plots and heatmaps were generated in Prism 9.3.1 (Graphpad) based on data exported from OMIQ. Volcano plots and correlation heatmaps were generated in R (R version 3.6.2; R Studio 1.2.5) with the packages “ggplot2” and “corrplot”, respectively. Microsoft Excel was used for some basic data maneuvering.

### In vitro stimulation and flow cytometry staining

After overnight resting, cells were stimulated in vitro in TexMACS™ Medium (Miltenyi Biotec) with 30 nM PMA (Sigma-Aldrich), 1 μM Ionomycin (Sigma-Aldrich) in presence of Brefeldin A and Monensin (Invitrogen) for 4 h.

Incubation was stopped by washing the cells in PBS and viable cells were detected with the Zombie NIR™ Fixable Viability Kit (Biolegend). Cells were, then, incubated with Human TrueStain FcX^TM^ (Biolegend) for quenching unspecific antibody staining. Cell surface staining was performed on ice with the following antibodies: PECy7-conjugated CD56 (HCD56, Biolegend) and TIGIT (VSTM3, Biolegend); APC-conjugated CD4 (RPA-T4, Biolegend) and HLA-DR (Tϋ39, Biolegend); Pacific-Blue-conjugated CD3ɛ (UCHT1, Biolegend); BV510-conjugated CD4 (OKT4, Biolegend). Next, cells were fixed and permeabilized with the Foxp3/Transcription Factor Staining Buffer Set (Invitrogen) according to the manufacturer’s instructions, and intracellular staining was performed with the following antibodies: FITC-conjugated FOXP3 (206D, Biolegend); PE-conjugated IL-10 (JES3-19F1, Biolegend); PerCP-conjugated IFN-γ (4S.B3, BioLegend) and BV510-conjugated Granzyme B (GB11, BD Horizon™). For assessment of degranulation capacity, PE-conjugated CD107a (H4A3, Biolegend) antibody was added during the 4 hours of in vitro stimulation. Data were acquired on a FACSCanto II (BD Biosciences) flow cytometer and analysed with FlowJo software (BD, Tree Star).

### In vitro CD16^+^CCR6^+^ T-cell differentiation

Freshly isolated PBMC from healthy donors’ blood were resuspended in MACS buffer (PBS with 0.5 % BSA, 2 mM EDTA) for enrichment of CD3^+^ T cells using MojoSort™ Human CD3 T Cell Isolation Kit (BioLegend) according to the manufacturer’s protocol. CD3^+^ T cells (0.2 x 10^6^ cells/well) were activated in round-bottom 96 multi-well plates (pre-coated with 5 μg/ml plate-bound Ultra-LEAF^TM^ anti-human CD3 (OKT3, Biolegend)) with soluble 1 μg/ml Ultra-LEAF^TM^ anti-human CD28 (CD28.2, Biolegend) and 20 IU/ml IL-2 (Preprotech) in complete TexMACS medium, in the presence of 10 % normal blood plasma (from human male AB plasma, Sigma) or CD or UC patients’ plasma (previously collected from the second cohort of patients and cryopreserved at −80°C until use). After 3 or 7 days, cells were harvested for flow cytometric staining as described in the previous section with the following antibodies for surface markers: FITC-conjugated CD16 (3G8, BioLegend); PECy7-conjugated CCR6 (G034E3, BioLegend); APC-conjugated CD8 (HIT8a, BioLegend); Pacific-Blue-conjugated CD3ɛ (UCHT1, BioLegend) and BV510-conjugated CD4 (OKT4, BioLegend).

### Statistical analysis

For Volcano plots: significance of Log2FoldChange was calculated in R (R version 3.6.1; R Studio 1.2.5) with Wilcoxon test and Benjamini Hochberg FDR correction for paired samples, and with Mann-Whitney test and Benjamini-Hochberg FDR correction for unpaired samples. Significant corrected p-value was set to p < 0.05 and Log2FoldChange threshold set to < −0.6 and > 0.6. Median marker intensity heatmaps were generated in Prism 9 (GraphPad) based on a raw clustered heatmap matrix generated and exported from OMIQ.ai (median intensity measure of markers). Bar-graphs for comparison of different population between groups (mass and flow cytometry) were generated in Prism 9 (GraphPad) and statistical testing were performed using: two-tailed Wilcoxon test for paired samples; two-tailed Mann-Whitney test for unpaired samples. P-value lower than p < 0.05 were considered significant. Correlation plots were generated in Prism 9 (GraphPad) with Spearman’s correlation test and data tested for linear regression.

### Study approval

Ethical approval was given by the ethical board of the University of Wuerzburg (proposal numbers 113/13, 46/11, 42/16), and all participants provided written, informed consent prior to participation.

## Supporting information

Combined Supplemental material

## Data Availability

All data produced are available online at flowrepository

https://flowrepository.org/id/FR-FCM-Z5DH

## Data Availability Statement

The data that support the findings of this study are available in https://flowrepository.org/ (FR-FCM-Z5DH).

## Author contributions

C.M.C. designed and performed research, analyzed and discussed the data thoroughly; mass cytometry was conducted by C.M.C. and A.R.S. at the DRFZ Berlin; M.M. collected the patients’ intestinal and blood samples. S.R.-H. graded patients’ tissues samples. A.J.R.O. organized CyTOF panels and software within the consortium; M.L. guided the consortium and discussed the study; A.R. provided financial support and discussed the data; H.-D.C. took part in the overall study design by mass cytometry and supported the data analyses; N.S. was decisive in the clinical part of the study design and discussed the data; H.E.M supervised and took part in the CyTOF analyses; F.B.-S. conceptualized the research goals, acquired major funding, designed the study, discussed the data, and wrote the manuscript together with C.M.C. All authors reviewed and approve of the manuscript.

## Acknowledgments

This work was mainly supported by the Else Kröner-Fresenius Foundation 2015_A232. Additional funding was received by the Wilhelm Sander-Foundation/2012.047.2 (F.B.-S.), the Deutsche Forschungsgemeinschaft (DFG, German Research Foundation), project number 324392634 - TRR 221 and FOR 2830 as well as the Fritz Thyssen Stiftung (Az. 10.13.2.215 and 10.17.2.012MN to F.B.-S.).

We are indebted to Heike Hirseland for excellent experimental assistance, Dr. Sabine Baumgart for mass cytometry support and Sebastian Ferrara for some help in bioinformatics. We thank Martina Prelog and Zeinab Mokhtari for valuable discussions and Lorenz Eing for his help with the graphical abstract.

