## Supplementary material for "Pro-inflammatory innate-like T cells are expanded in the blood and inflamed intestine in Crohn’s Disease": Combined Supplemental material

### Supplemental Table 1. Patients of cohort 1 / CyTOF

| Patient ID | Disease | Sex | Age range (years) | Intestinal region resected | Disease history range (years) | Previous medication | Current medication | Malignancy | Pathological score | CRP preoperative (mg/L) | Leucocyte count preoperative ($10^9$cells/L) |
|---|---|---|---|---|---|---|---|---|---|---|---|
| PT1 | Crohn's Disease | F | 51 - 60 | Terminal Ileum | 31 - 40 | Corticosteroids Adalimumab | NA | None | Severe acute | 0.41 | 11 |
| PT2 | | M | 31 - 40 | | 11 - 20 | Purine analog Corticosteroids | Adalimumab | None | Severe acute | 0.11 | 9 |
| PT3 | | F | 61 - 70 | | 31 - 40 | None | None | None | Severe acute | 0.02 | 3.4 |
| PT4 | | F | 21 - 30 | | 0 - 10 | 5-ASA Corticosteroids Purine analogs Adalimumab Methotrexate | Corticosteroids Adalimumab | None | Moderate acute | 3.32 | 6.9 |
| PT5 | | M | 21 – 30 | | 0 - 10 | 5-ASA | Corticosteroids | None | Severe acute | 0.89 | 9.5 |
| PT6 | | M | 41 - 50 | | 0 - 10 | Cortocosteroids | Corticosteroids | None | Chronic inflammation | 5.18 | 11.4 |
| PT7 | | M | 21 -30 | | 0 - 10 | Purine analogs Vedolizumab Corticosteroids | Vedolizumab | None | Severe acute | 0.35 | 8.3 |
| PT8 | | F | 41 -50 | | 11 - 20 | Infliximab | Infliximab | None | Mild acute | 1.54 | 8.6 |
| PT9 | | F | 31 - 40 | | 0 - 10 | NA | Corticosteroids | None | Severe acute | 0.18 | 7.9 |
| PT10 | | F | 31 -40 | | 0 - 10 | Adalimumab Purine analogs Corticosteroids | Infliximab | None | Moderate acute | 2.78 | 8.7 |
| PT11 | | M | 31 - 40 | | 0 - 10 | Corticosteroids 5-ASA | Corticosteroids | Yes | Moderate acute | 1.7 | 8.8 |
| PT12 | | M | 21 -30 | | 0 - 10 | Corticosteroids | Inflixmab | None | Chronic inflammation | 0.32 | 4.9 |
| PT13 | | M | 31 - 40 | | 0 - 10 | Purine analogs | NA | None | Moderate inflammation | 0.24 | 9.7 |
| PT14 | | F | 31 - 40 | | 11 - 20 | Infliximab Adalimumab | Vedolizumab VitD | None | Chronic inflammation | 0.42 | 8.3 |
| PT15 | | M | 21 - 30 | | 0 - 10 | Corticosteroids Pantoprazole | NA | None | Chronic inflammation | 1.17 | 6.0 |
| PT16 | | M | 51 - 60 | | 0 - 10 | Infliximab Metroprolol | NA | None | Moderate acute | 8.83 | 6.9 |
| PT17 | | M | 61 - 70 | | 0 - 10 | Infliximab Corticosteroids | NA | None | Mild acute | 0.50 | 10.0 |

### Supplemental Table 2. Patients of cohort 2 / fluorescence-based flow cytometry

| Patient ID | Disease | Sex | Age range (years) | Intestinal region resected | Disease history range (years) | Previous medication | Current medication | Malignancy | CRP preoperative (mg/L) | Leucocyte count preoperative ($10^9$cells/L) |
|---|---|---|---|---|---|---|---|---|---|---|
| PT18 | Crohn's Disease | M | 21 - 30 | Terminal Ileum | 0 - 10 | Corticosteroids, Infliximab, Purine analogs | Ustekinumab | None | 2.17 | 6.0 |
| PT19 | | M | 61 - 70 | | 11 - 20 | Adalimumab | | Yes | 5.25 | 10.3 |
| PT20 | | F | 21 - 30 | | 0 - 10 | Purine analog, Adalimumab | | None | 3.75 | 5.6 |
| PT21 | | M | 41 - 50 | | 11 - 20 | 5-ASA, Corticosteroids, Infiliximab, Vedolizumab, Ustekinumab | Adalimumab | None | 0.11 | 10.9 |
| PT22 | | M | 21 - 30 | | 0 - 10 | Corticosteroids, Adalimumab, Ustekinumab | | None | 0.55 | 7.6 |
| PT23 | | F | 51 - 60 | | 31 - 40 | Corticosteroids, Purine analogs | Infliximab | None | 0.13 | 7.3 |
| PT24 | | F | 61 - 70 | | 31 - 40 | Corticosteroids, Vedolizumab, Infliximab, Adalimumab, Purine analogs, Golimumab, Certolizumab | Ustekinumab | None | <0.1 | 9.1 |
| PT25 | | M | 61 -70 | | 21 - 30 | Corticosteroids, VitB12, VitD | NA | None | 1.38 | 10.3 |
| PT26 | | F | 21 - 30 | | 11 - 20 | Purin analogs, Adalimumab | NA | Yes | <0.1 | 6.3 |
| PT27 | Ulcerative Colitis | M | 21 - 30 | Colon | 0 - 10 | Adalimumab, Ustekinumab, Purine analog, Corticosteroids | NA | None | 0.67 | 7.5 |
| PT28 | | F | 41 - 50 | | 0 - 10 | Adalimumab, Ustekinumab, Corticosteroids, Vedolizumab | Corticosteroids VitD | None | 1.23 | 8.3 |
| PT29 | | F | 61 - 70 | | 0 - 10 | Corticosteroids, Infliximab, Golimumab, Vedolizumab, Ustekinumab | Tofacitinib | None | 1.91 | 7.0 |

### Supplemental Table 3. Mass cytometry antibody panel

| Metal isotope | Target | Alias | Antibody clone | Supplier | Staining |
|---|---|---|---|---|---|
| **89Y** | B2M | | 2M2 | Biolegend | barcode |
| **104Pd** | B2M | | 2M2 | Biolegend | barcode |
| **105Pd** | CD57 | HNK1 | HCD57 | Biolegend | surface |
| **108Pd** | B2M | | 2M2 | Biolegend | barcode |
| **110Pd** | B2M | | 2M2 | Biolegend | barcode |
| **113In** | HLA-DR | MHC-II | L243 | DRFZ | surface |
| **115In** | CD3ε | | UCHT1 | DRFZ | surface |
| **140Ce** | CD45 | PTPRC | Hi30 | Biolegend | surface |
| **141Pr** | CCR6 | CD196 | G034E3 | Fluidigm | surface |
| **142Nd** | CD27 | TNFRSF7 | 2E4 | Sanquin | surface |
| **143Nd** | CD127 | IL-7Rα | A019D5 | Fluidigm | surface |
| **144Nd** | CD4 | | RPA-T4 | Biolegend | surface |
| **145Nd** | CXCR5 | CD185 | REA103 | Miltenyi Biotec | surface |
| **146Nd** | TCR Va7.2 | | 3C10 | BIolegend | surface |
| **147Sm** | PD-1 | CD279 | EH12.2H7 | Biolegend | surface |
| **148Nd** | α4β7 integrin | | Vedolizumab | Takeda | surface |
| **149Sm** | CD25 | IL-2Rα | 2A3 | Fluidigm | surface |
| **150Nd** | NKp46 | CD335 | 9E2 | Miltenyi Biotec | surface |
| **151Eu** | ICOS | CD278 | C398.4A | Fluidigm | surface |
| **152Sm** | CD103 | αE integrin | Ber-ACT8 | Biolegend | surface |
| **153Eu** | TIGIT | Vstm3 | MBSA43 | Fluidigm | surface |
| **154Sm** | TIM-3 | CD366 | F38-2E2 | Fluidigm | surface |
| **155Gd** | IRF4 | | 3E4 | Fluidigm | intracellular 1 |
| **156Gd** | CXCR3 | CD183 | G025H7 | Fluidigm | surface |
| **157Gd** | anti-PE | | PE001 | Biolegend | surface |
| **158Gd** | CCR4 | CD194 | L291H4 | Fluidigm | surface |
| **159Tb** | CCR7 | CD197 | G043H7 | Biolegend | surface |
| **160Gd** | TBET | | 4B10 | Fluidigm | intracellular 1 |
| **161Dy** | CD28 | | L293 | BD Biosciences | surface |
| **162Dy** | FOXP3 | | PCH101 | Fluidigm | intracellular 1 |
| **163Dy** | CRTH2 | CD294 | BM16 | Fluidigm | surface |
| **164Dy** | CCR10 | GPR-2 | 314305 | R&D Systems | surface |
| **165Ho** | LAG-3 | CD223 | 11C3C65 | Fluidigm | surface |
| **166Dy** | NKp44 | CD336 | 2.29 | Miltenyi Biotec | surface |
| **167Er** | GATA-3 | | REA174 | Miltenyi Biotec | intracellular 1 |
| **168Er** | RORγt | | REA278 | Miltenyi Biotec | surface |
| **169Tm** | TCRγδ | | 11F2 | Miltenyi Biotec | surface |
| **170Er** | CTLA-4 | CD152 | 14D3 | Fluidiqm | surface |
| **171Yb** | CD161 | KLRB1 | HP-3G10 | Biolegend | surface |
| **172Yb** | Anti-Cy5/AF647 | | AM1-7H6.16.44 | Miltenyi Biotec | intracellular 2 |
| **173Yb** | GranzymeB | CTLA1 | GB11 | Fluidigm | intracellular 1 |
| **174Yb** | CD45RA | | 4G11 | DRFZ | surface |
| **176Yb** | CD56 | NCAM-1 | NCAM16.2 | Fluidigm | surface |
| **194Pt** | CD8α | | GN11/134D7 | DRFZ | surface |
| **195Pt** | B2M | | 2M2 | Biolegend | barcode |
| **196Pt** | B2M | | 2M2 | Biolegend | barcode |
| **198Pt** | B2M | | 2M2 | Biolegend | barcode |
| **209Bi** | CD16 | FcγRIII | 3G8 | Fluidigm | surface |
| **PE** | CD19 | | LT19 | Miltenyi Biotec | dump(surface) |
| **PE** | CD20 | | LT20 | Miltenyi Biotec | dump(surface) |
| **PE** | CD36 | Platelet glycoprotein 4 | AC106 | Miltenyi Biotec | dump(surface) |
| **PE** | CD123 | IL-3Rα | AC145 | Miltenyi Biotec | dump(surface) |
| **AF647** | IRF1 | | D5E4 | Cell Signaling Technology | intracellular 1 |

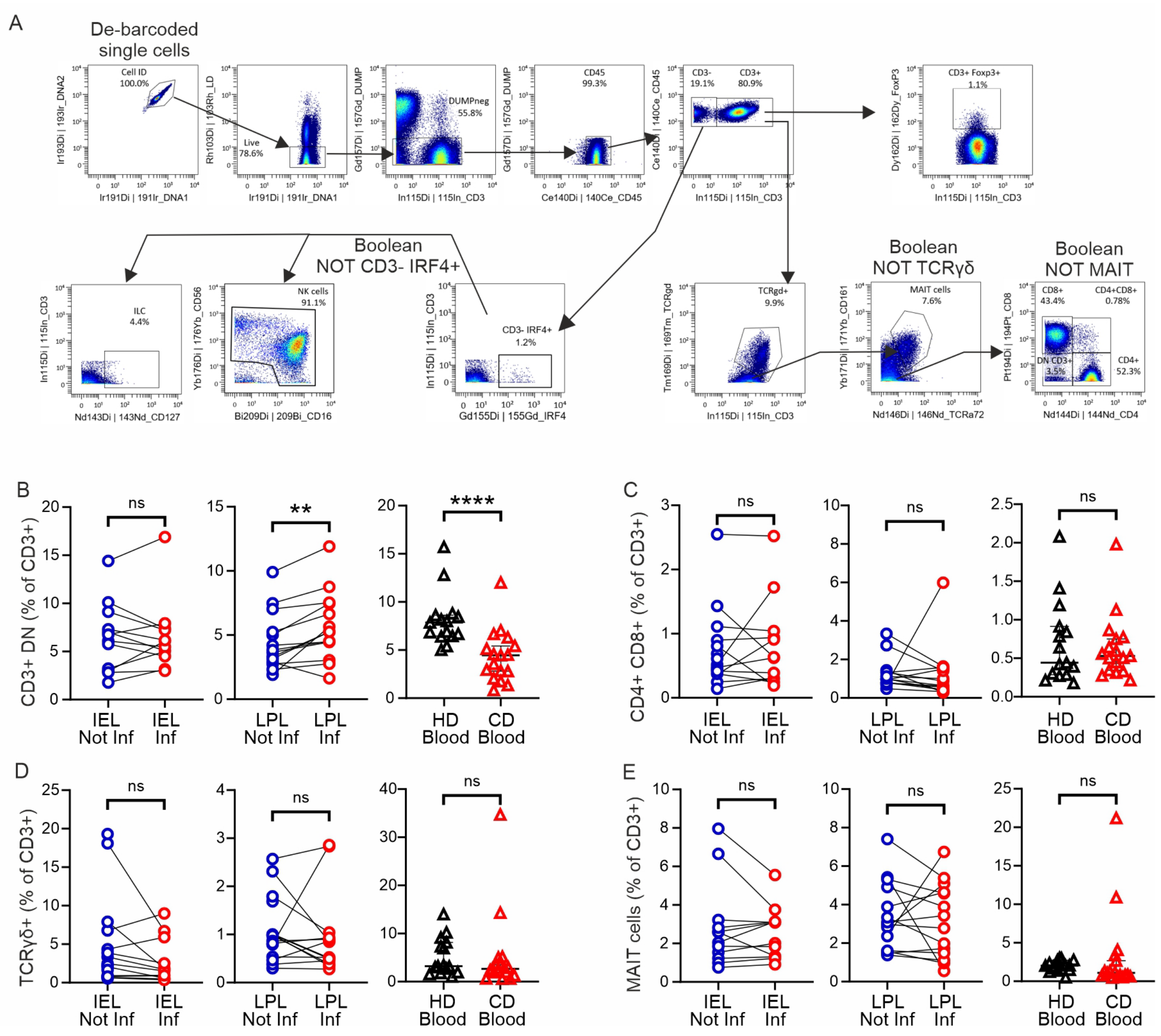


**Figure S1. Frequencies of CD3⁺CD4⁻CD8⁻ T cells enriched in inflamed LP, but were less among PBMC of CD patients.** (**A**) Gating strategy (starting from single cells of de-barcoded samples) used for identification of all the different CD3⁺ and CD3⁻ cells populations analysed in this study. The gating was performed in OMIQ. (**B**) Frequencies of CD3⁺CD4⁻CD8⁻ T cells, (**C**) CD3⁺CD4⁺CD8⁺ T cells, (**D**) CD3⁺TCRγδ⁺ T cells and (**E**) CD3⁺MAIT cells between inflamed and not inflamed epithelium and LP as well as CD and HD blood. Statistical significances were calculated by two-tailed Wilcoxon test for paired intestinal samples and by two-tailed Mann-Whitney test for not paired blood samples. (**, $p<0.01$; ****, $p<0.0001$).

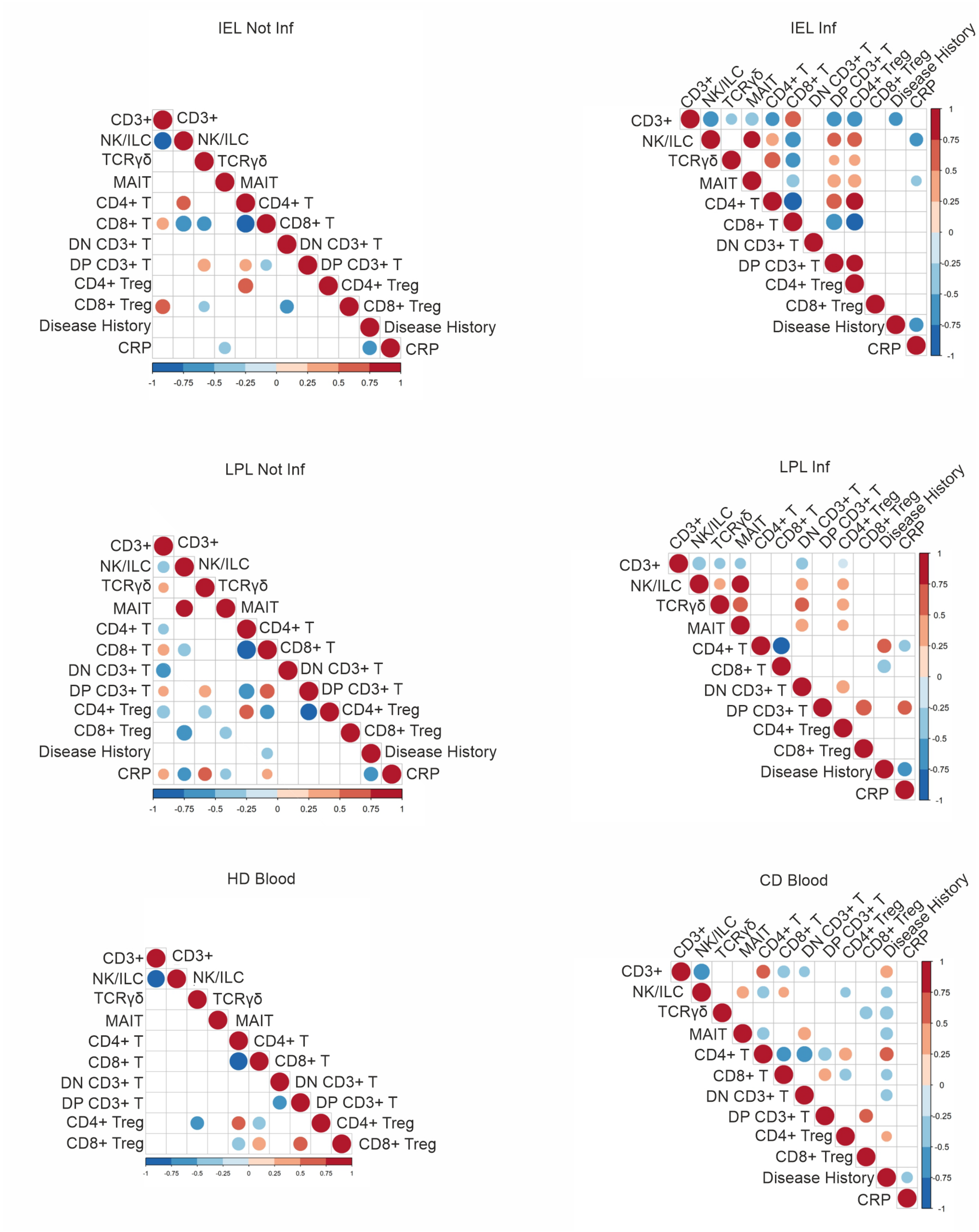


**Figure S2. Altered correlation of main cell populations under inflammation in CD patients.** Correlation heatmaps of main lymphocyte populations and patients clinical data in intestinal epithelium (top), lamina propria (middle) and blood (bottom). Color and dot size indicate correlation coefficient. Only significant correlations are shown. Spearman’s correlation test, $p < 0.05$.

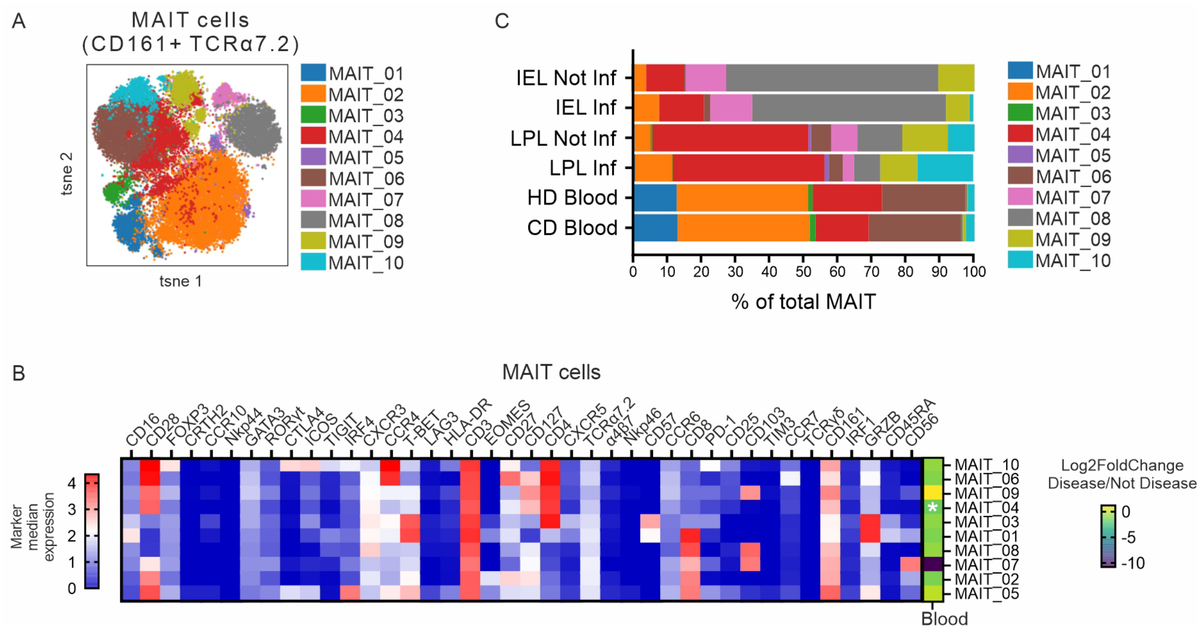


**Figure S3. Identification of 10 MAIT-cell clusters.** MAIT cells were gated as shown in Supplemental Figure 1A. (**A**) tsne projection of MAIT cells from all blood and tissue samples. 10 MAIT-cell clusters derived from FlowSOM clustering are indicated. (B) Heatmap (calculated from all samples) of the 10 MAIT cell clusters (rows) shows median arcsinh-transformed signal intensities of 40 markers (columns) used for cluster analysis. The right part of the heatmap displays differences in abundance for each cluster (as % of $CD3^{+}$ T cells) between CD and HD blood, as Log2FoldChange. Asterisks (*) indicate clusters of significantly different abundance between conditions. Statistics were calculated with Mann-Whitney test with FDR correction. White asterisk: cluster significantly less abundant in disease status (FDR corrected p value < 0.05, Log2FoldChange < -0.6). (**C**) Abundance of each MAIT-cell cluster (as percentage) within the total MAIT cells in each tissue and condition.

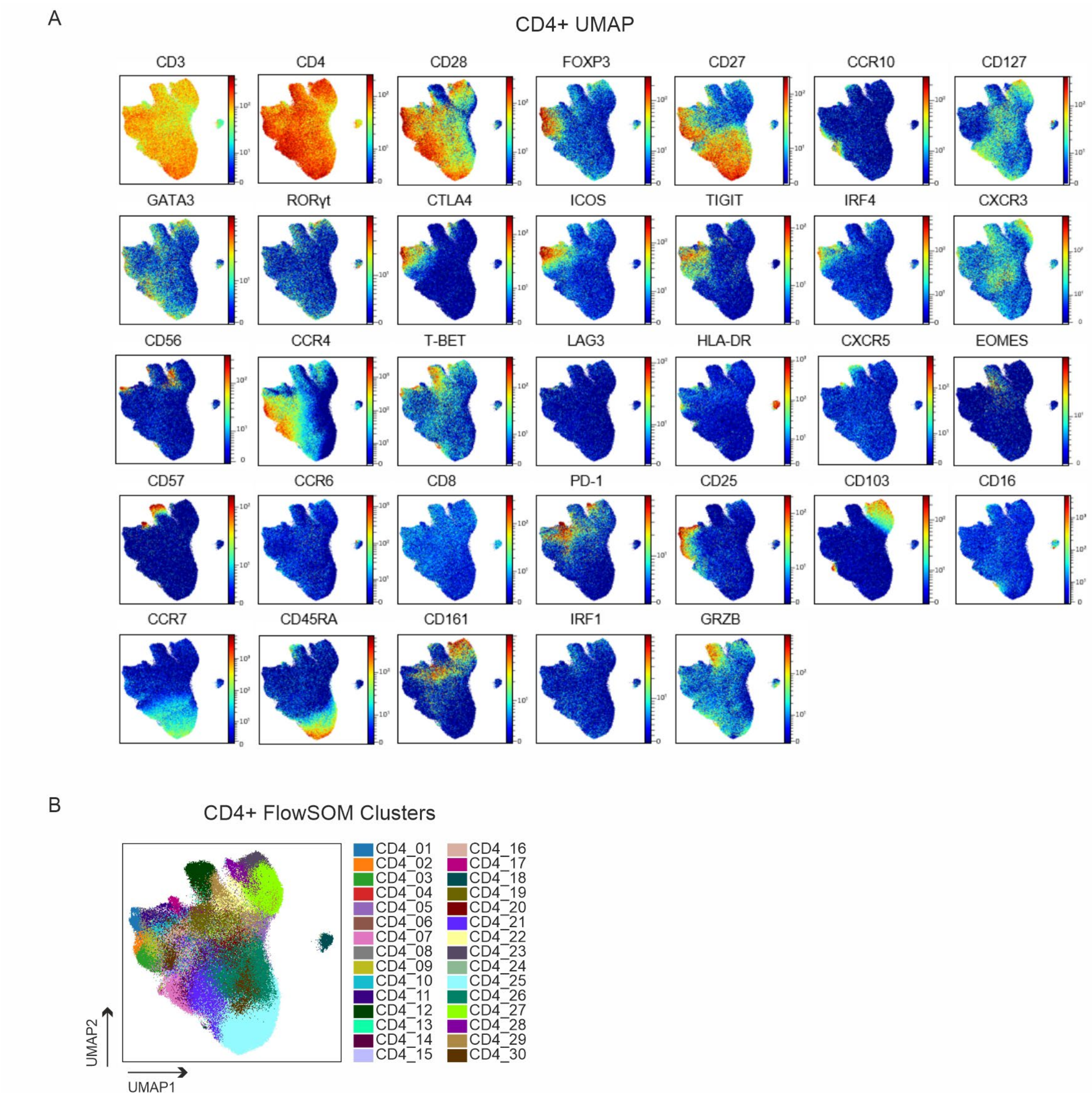


**Figure S4. CD4⁺ T-cell analysis.** (**A**) UMAP plots, from all the intestinal and blood samples, show expression in CD4⁺ T cells of selected markers. (**B**) UMAP plot, from all intestinal and blood samples, depicts distribution of 30 CD4⁺ T-cells clusters identified with FlowSOM.

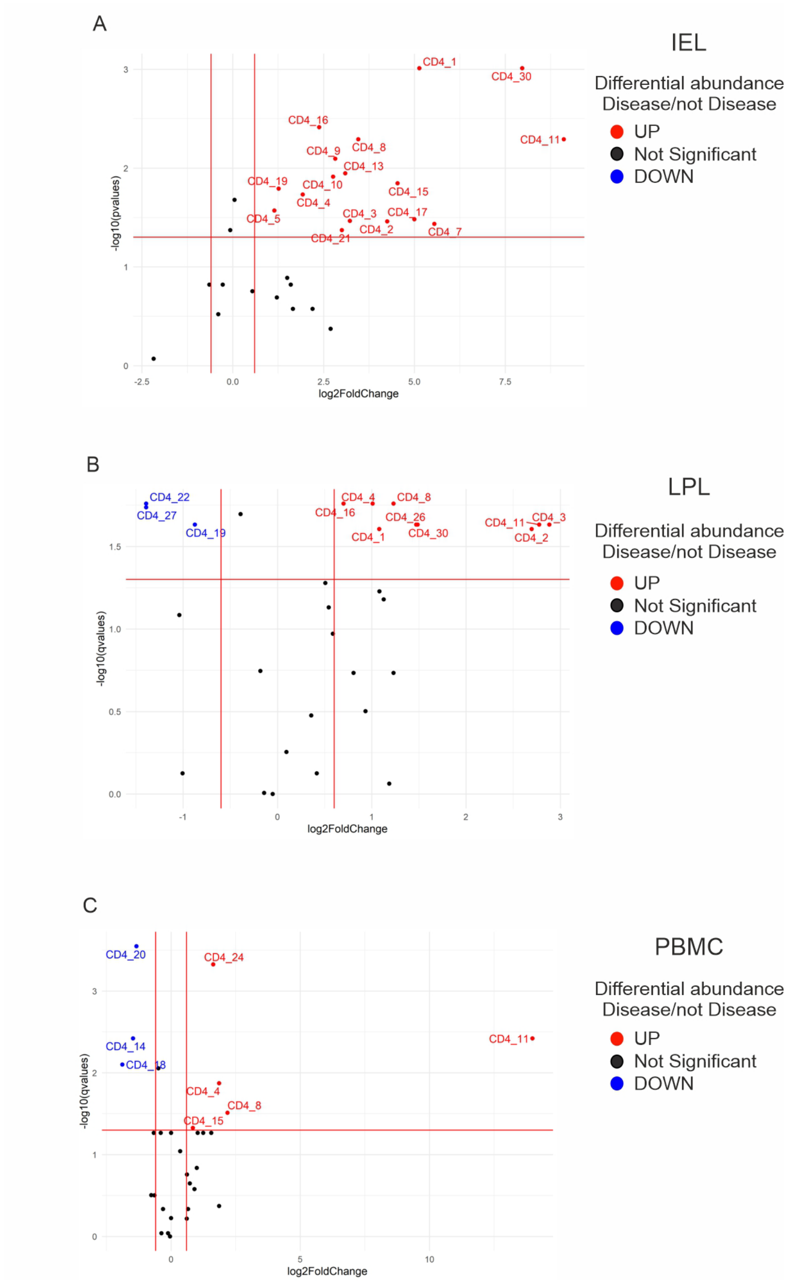


**Figure S5. Volcano plots indicate overall enriched CD4$^{+}$ T-cell subpopulations in inflamed intestinal tissue of CD patients.** (**A**) Volcano plot showing Log2FoldChange of IEL Inf on IEL Not Inf of all CD4$^{+}$ T-cell clusters identified by FlowSOM algorithm. Wilcoxon test for paired intestinal samples. Cut off for significant changes: FDR corrected p value $< 0.05$, Log2FoldChange $< -0.6$ and $> 0.6$. (**B**) Volcano plot showing Log2FoldChange of LPL Inf on LPL Not Inf of all CD4$^{+}$ T-cell clusters identified by FlowSOM algorithm. Wilcoxon test for paired intestinal samples. Cut off for significant changes: FDR corrected p value $< 0.05$, Log2FoldChange $< -0.6$ and $> 0.6$. (**C**) Volcano plot showing Log2FoldChange of CD blood on HD blood of all CD4$^{+}$ T cells clusters identified by FlowSOM algorithm. Mann-Whitney test for not paired blood samples. Cut off for significant changes: FDR corrected p value $< 0.05$, Log2FoldChange $< -0.6$ and $> 0.6$.

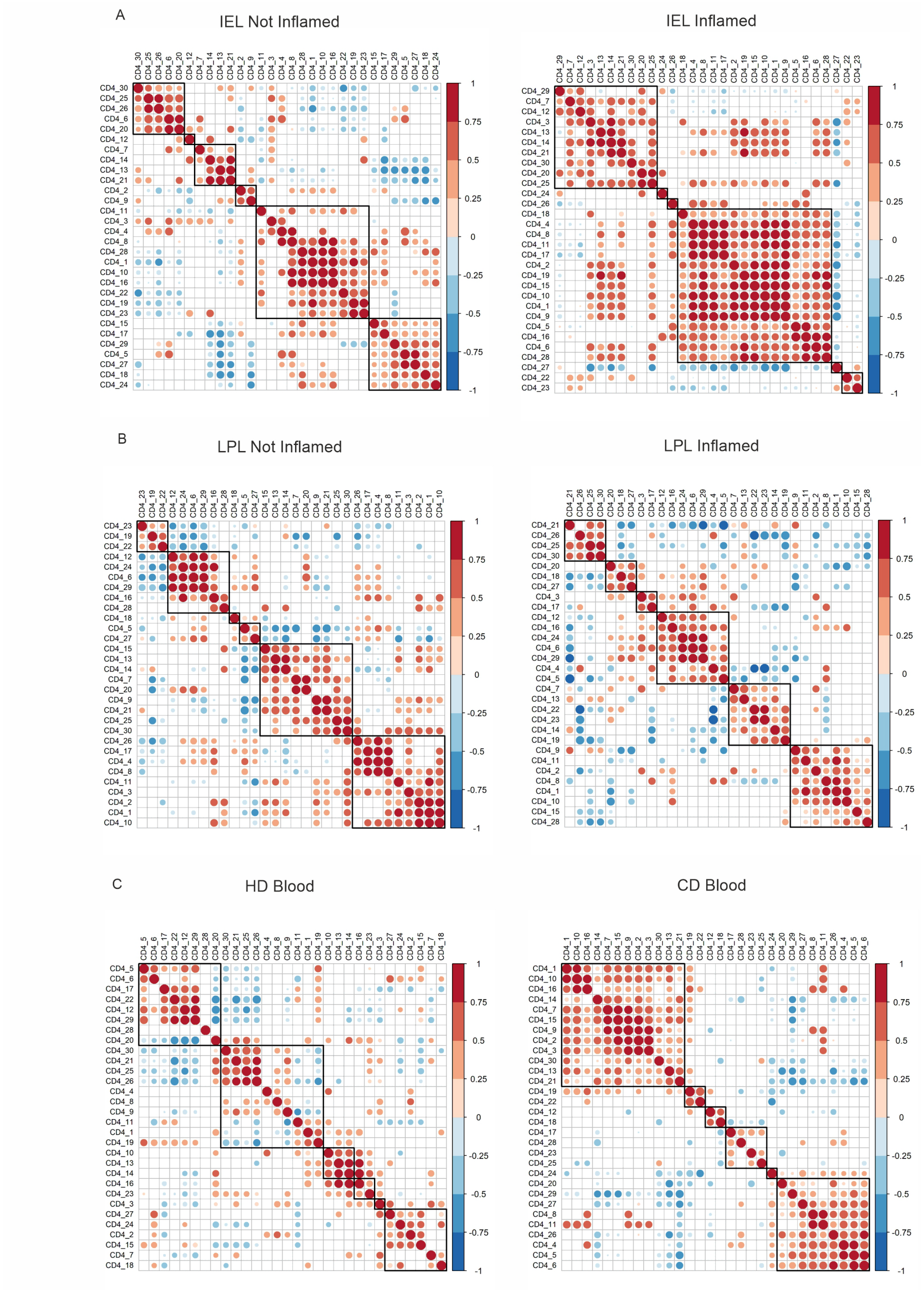


**Figure S6. Inflammation in CD patients disturbed CD4$^+$ T-cell subpopulation correlations.** (**A**) Correlation heatmaps of CD4$^+$ T-cell clusters in IEL Not Inf (left) and IEL Inf (right). (**B**) Correlation heatmaps of CD4$^+$ T-cell clusters in LPL Not Inf (left) and LPL Inf (right). (**C**) Correlation heatmaps of CD4$^+$ T-cell clusters in HD blood (left) and CD blood (right). (A, B, C) Color and dot size indicate correlation coefficient. Only significant correlations are shown. Spearman's correlation test, $p < 0.05$.

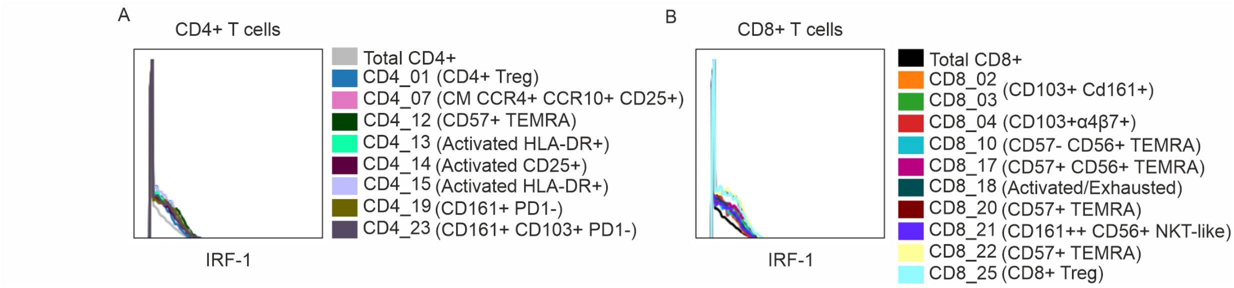


**Figure S7. IRF1 expression correlated with activation, $T_{EMRA}$ differentiation and innate-like phenotype in T cells.** Overlay histograms show the expression of IRF-1 in defined $CD4^+$ (**A**) and $CD8^+$ (**B**) T-cell clusters.

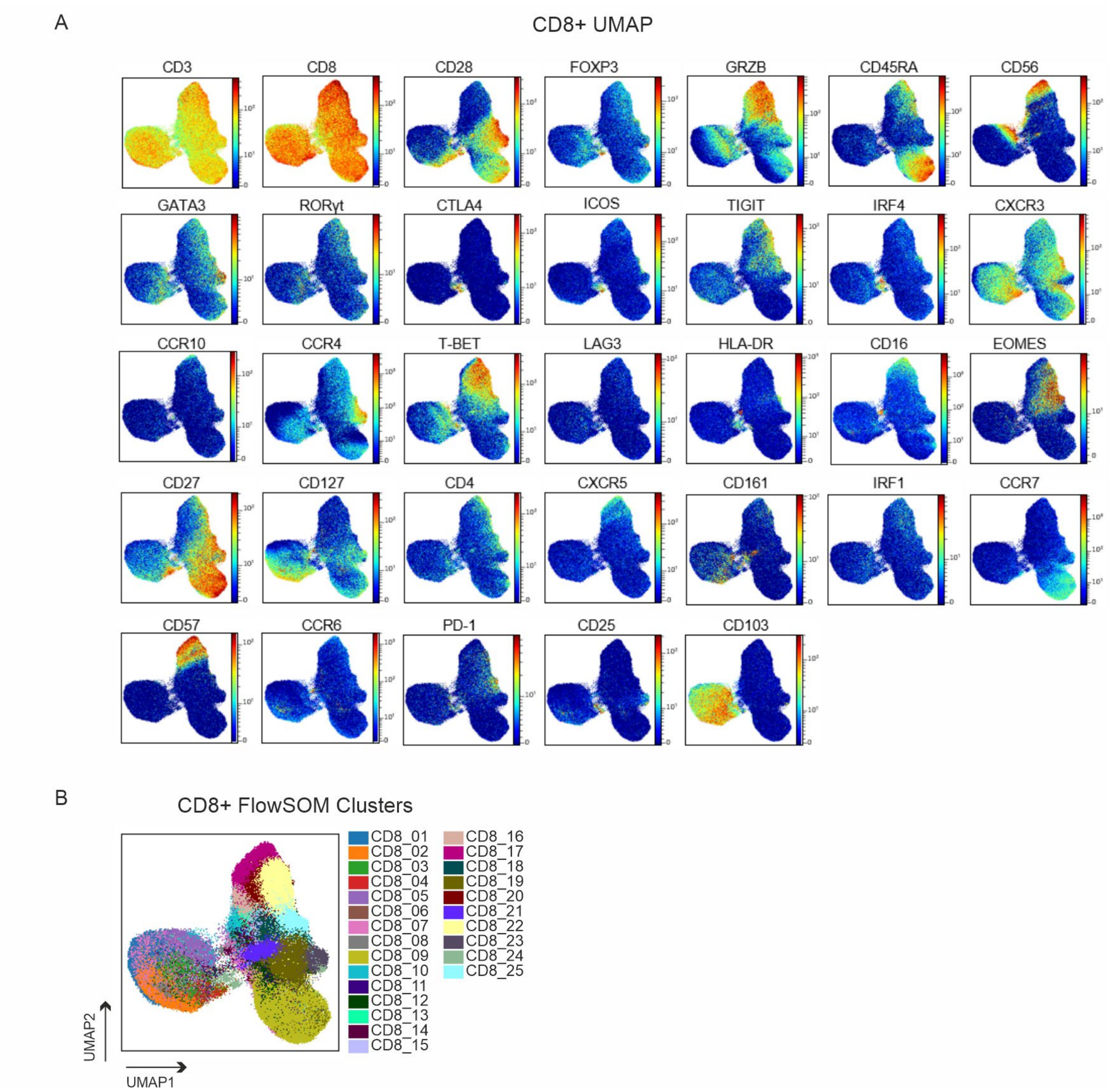


**Figure S8. CD8$^+$ T-cell analysis.** (**A**) UMAP plots, from all the intestinal and blood samples, show expression in CD8$^+$ T cells of selected markers. (**B**) UMAP plot, from all intestinal and blood samples, depicts distribution of 25 CD8$^+$ T-cell clusters identified with FlowSOM.

Fig. S9

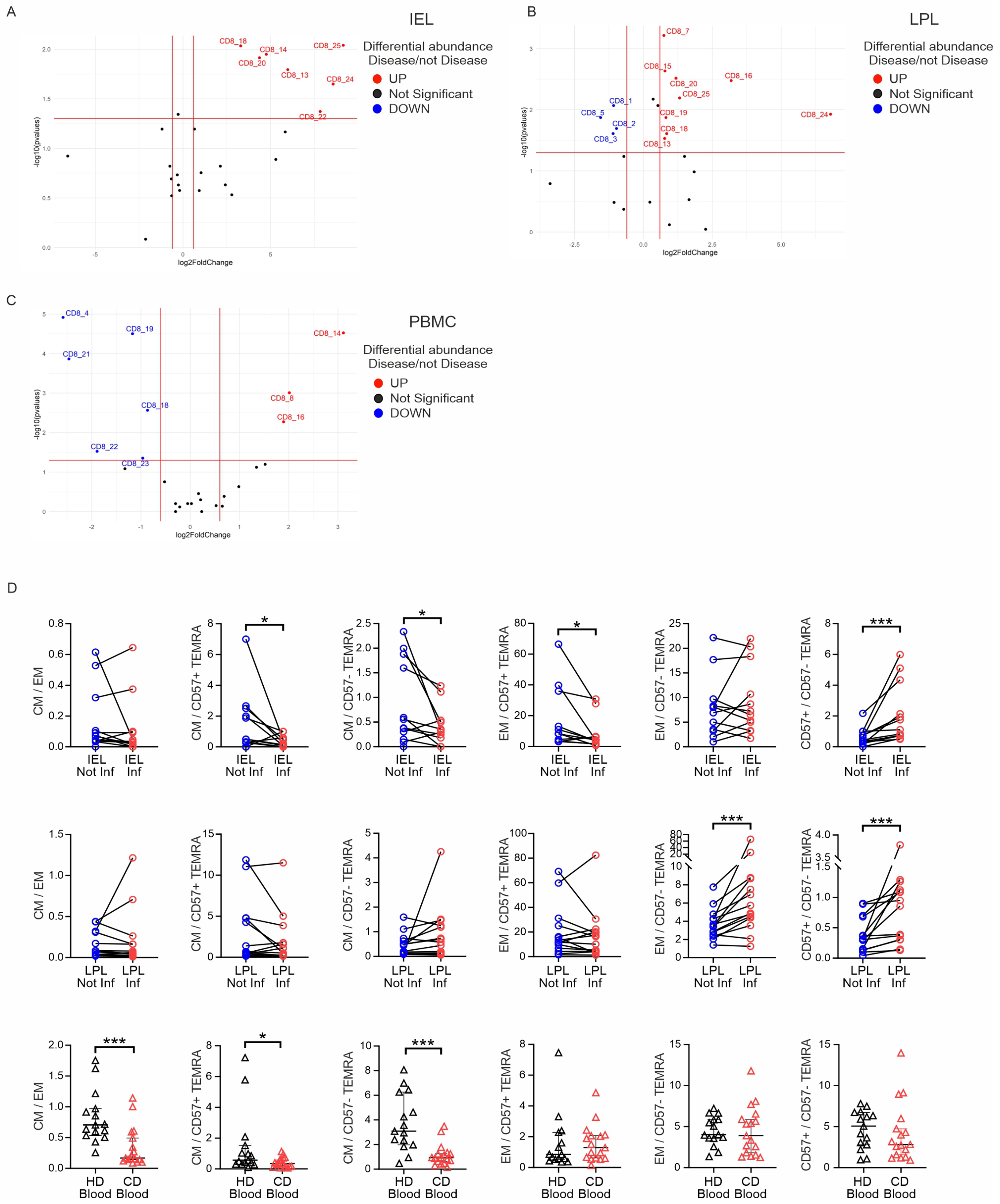


**Figure S9. Relative abundance of CD8⁺ $T_{CM}$ was reduced, while that of CD57⁺ $T_{EMRA}$ was enhanced upon inflammation in CD.** (**A**) Volcano plot showing Log2FoldChange of IEL Inf on IEL Not Inf of all CD4⁺ T-cell clusters identified by FlowSOM algorithm. Wilcoxon test for paired intestinal samples. Cut off for significant changes: FDR corrected p value < 0.05, Log2FoldChange < -0.6 and > 0.6. (**B**) Volcano plot showing Log2FoldChange of LPL Inf on LPL Not Inf of all CD4⁺ T-cell clusters identified by FlowSOM algorithm. Wilcoxon test for paired intestinal samples. Cut off for significant changes: FDR corrected p value < 0.05, Log2FoldChange < -0.6 and > 0.6. (**C**) Volcano plot showing Log2FoldChange of CD blood on HD blood of all CD4⁺ T-cell clusters identified by FlowSOM algorithm. Mann-Whitney test for not paired blood samples. Cut off for significant changes: FDR corrected p value < 0.05, Log2FoldChange < -0.6 and > 0.6. (**D**) Ratio of CD8⁺ $T_{CM}/T_{EM}$, $T_{CM}/T_{EMRA}$ and $T_{EM}/T_{EMRA}$ in all tissues. Two-tailed Wilcoxon test for paired intestinal samples and Mann-Whitney test for not paired blood samples. (*, $p < 0.05$; ***, $p < 0.001$).

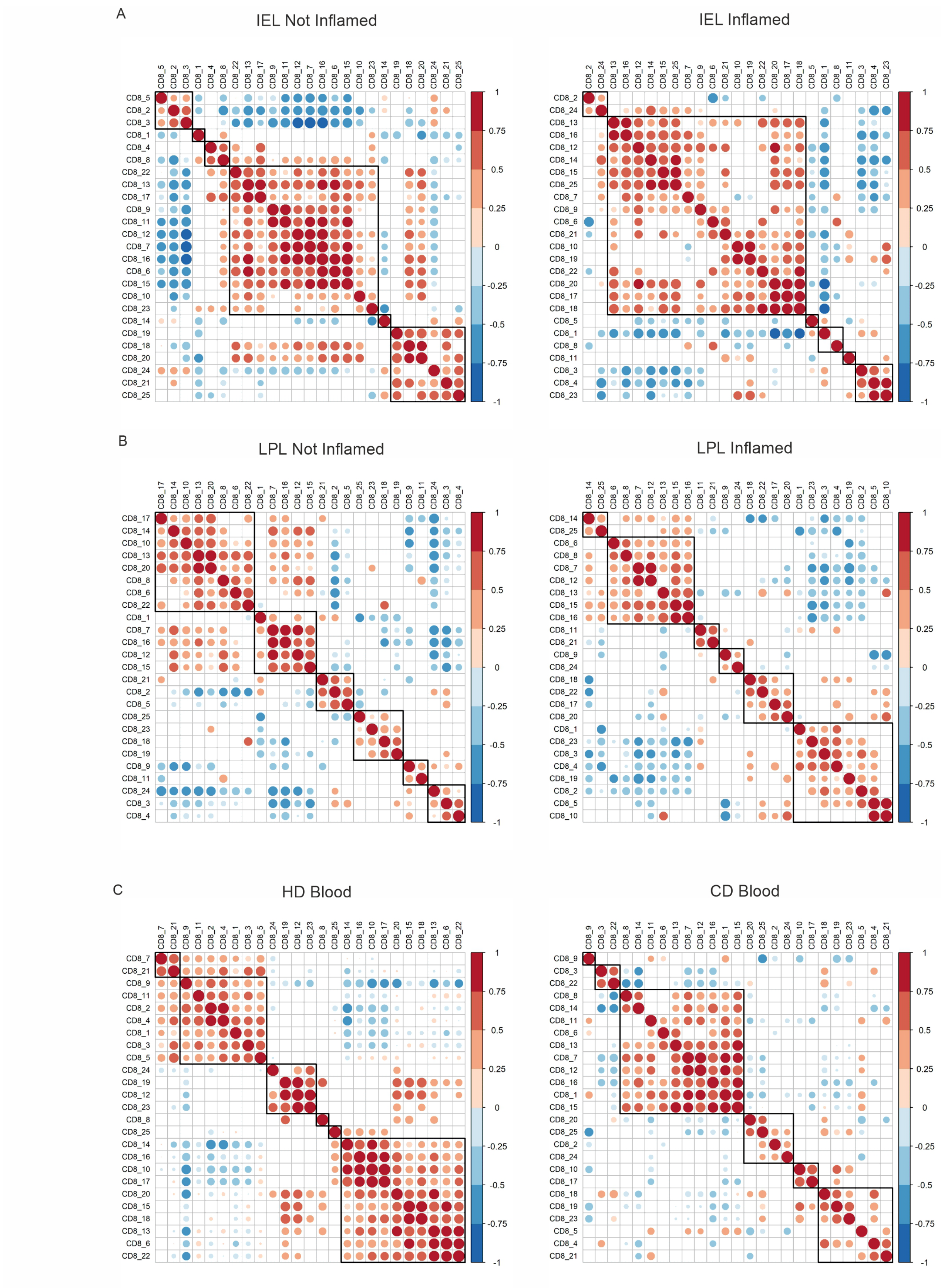


**Figure S10. Inflammation in CD patients disturbed CD8⁺ T-cell subpopulation correlations.** (**A**) Correlation heatmaps of CD8⁺ T-cell clusters in IEL Not Inf (left) and IEL Inf (right). (**B**) Correlation heatmaps of CD8⁺ T-cell clusters in LPL Not Inf (left) and LPL Inf (right). (**C**) Correlation heatmaps of CD8⁺ T-cell clusters in HD blood (left) and CD blood (right). (A, B, C) Color and dot size indicate correlation coefficient. Only significant correlations are shown. Spearman's correlation test, $p < 0.05$.

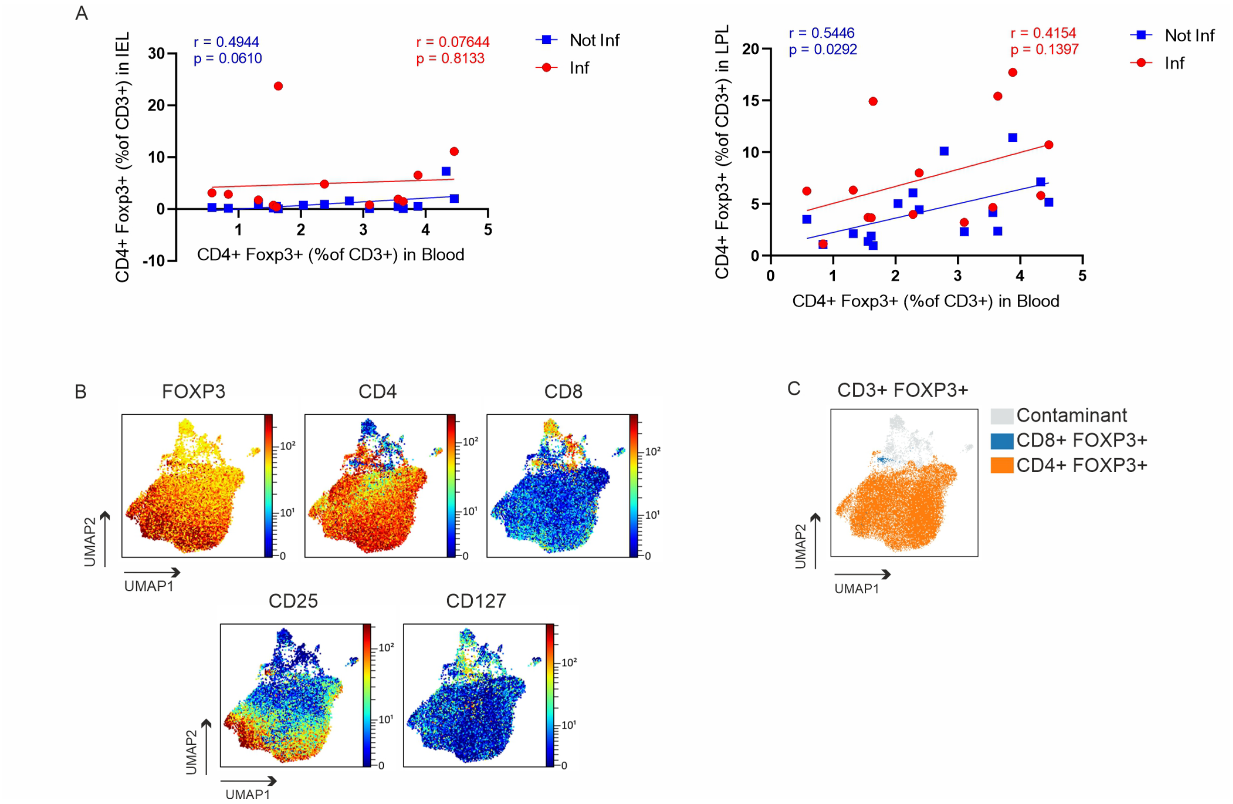


**Figure S11. Less Tregs among PBMCs did not correlate with more Tregs in CD patients' intestinal tissues.** (**A**) Correlation between frequency of CD4$^+$FOXP3$^+$ Treg cells in IEL (left) and in LPL (right) and frequencies of the same population in CD blood. Spearman's correlation test and linear regression. (**B**) Color continuous UMAP of CD3$^+$FOXP3$^+$ from all concatenated samples show expression levels (dark red, high expression; dark blue, no expression) of Treg-lineage markers. (**C**) Not real Treg (CD127$^+$) contaminating the manually gated CD3$^+$ FOXP3$^+$ T cells were excluded from the analysis. Overlaid UMAP plot shows not real Tregs, CD8$^+$ and CD4$^+$ Tregs.

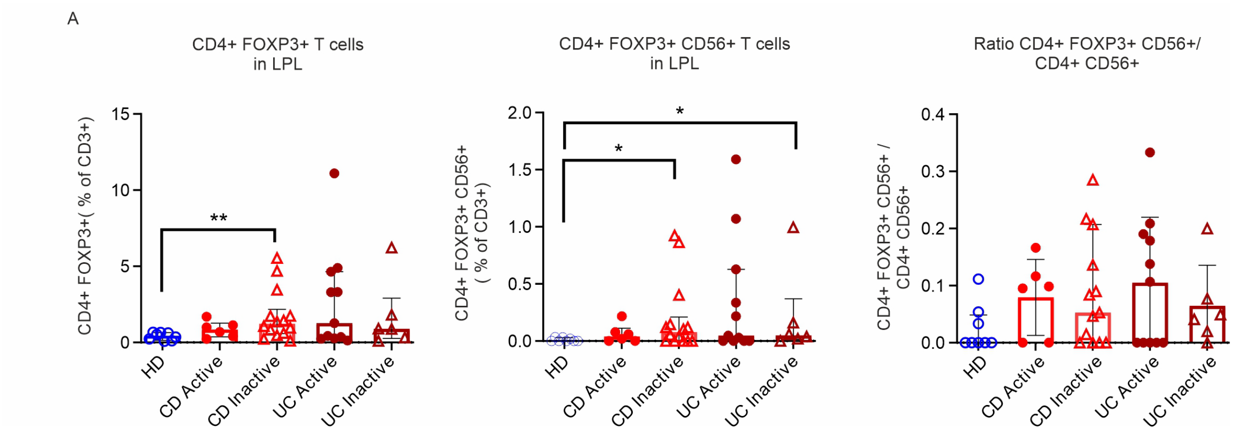


**Figure S12. Re-evaluation of published data confirms the presence of aberrant CD4⁺FOXP3⁺CD56⁺ T cells in IBD patients.** Analysis performed on FCS files shared on OMIQ repository by Mitsialis et al. 2020. (**A**) Frequencies of $CD4^{+}FOXP3^{+}$ T cells in LPL of healthy donors, CD active and not active, UC active and not active. (**B**) Frequencies of $CD4^{+}FOXP3^{+}CD56^{+}$ T cells in LPL of healthy donors, CD active and not active, UC active and not active. (**C**) Ratio of $CD4^{+}FOXP3^{+}CD56^{+}$ T cells vs. $CD56^{+}$ T cells in LPL of healthy donors, CD active and not active, UC active and not active. Two-tailed Mann-Whitney test. (*, $p < 0.05$; **, $p < 0.01$).

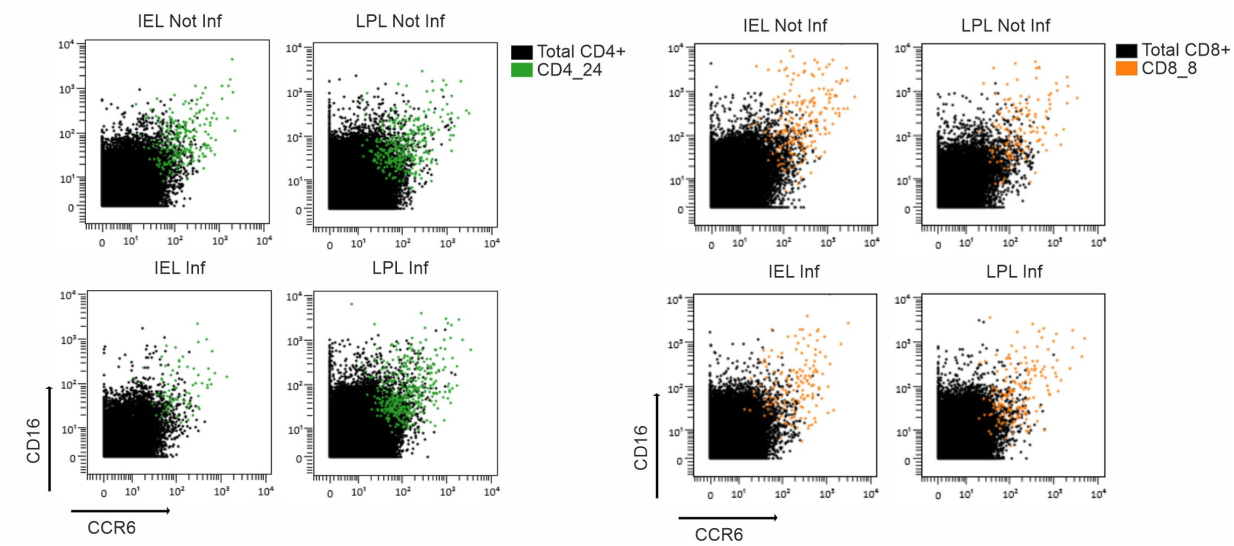


**Figure S13. CD16$^+$CCR6$^+$ T cells were present, irrespective of inflammation, in the CD patients' intestines.** Overlaid IEL and LPL (inflamed and not inflamed) samples in two-dimension dot plots show the expression of CD16 and CCR6 of the clusters CD4_24 and CD8_8.
